# Integrating dorsolateral prefrontal cortex multi-omics and GWAS summary data enhances gene-mapping of Parkinson’s disease

**DOI:** 10.64898/2026.05.23.26353951

**Authors:** Qiang Liu, Shinya Tasaki, David A. Bennett, Nicholas T. Seyfried, Philip L. De Jager, Vilas Menon, Aron S. Buchman, Jingjing Yang

## Abstract

Although dorsolateral prefrontal cortex (DLPFC) is not the main site of the Parkinson disease’s (PD) defining pathology, DLPFC is still functionally and structurally relevant as PD progresses - particularly for the cognitive/executive dimension of PD in later stages. To enhance the gene-mapping of PD, we integrated xQTL weights derived from bulk RNA-seq (n=931), single-nucleus RNA-seq (n=415), and bulk proteomics (n=716) data of DLPFC with the largest available GWAS summary data of PD in European population. Through integrative Omnibus TWAS and PWAS analyses, we detected risk genes associated with PD through genetically predicted bulk or cell-type-aware gene expression, or bulk protein abundances in DLPFC. We detected 39 significant risk genes by bulk TWAS, 66 unique genes by cell-type-aware TWAS across six brain cell types, and 17 by bulk PWAS. Importantly, 57.9% (11/19) bulk and 62.5% (20/32) cell-type-aware independent TWAS risk genes had PWAS locus support. Comparing these TWAS/PWAS findings in DLPFC to significant PD TWAS risk genes detected using the bulk RNA-seq data of substantia nigra from the GTEx V10 (n=183) and the same GWAS summary data of PD, 53.8% (21/39) bulk TWAS risk genes, 36.4% (24/66) unique cell-type-aware TWAS genes, and 17.6% (3/17) PWAS genes in DLPFC were also found significant in substantia nigra. Protein–protein interaction analyses indicated connectivity of our detected risk genes in DLPFC with established PD-associated genes such as *MAPT, SNCA*, and *LRRC37A*. Cell-type-aware TWAS risk genes were enriched in *apoptosis signaling*, and *T-cell activation* was enriched across the bulk TWAS, cell-type-aware TWAS, and PWAS results.

## Introduction

Parkinson’s disease (PD) is the second most common neurodegenerative disorder worldwide, and its incidence is rising in step with global population aging^1^. Recent large-scale genome-wide association studies (GWASs) have uncovered ∼90 independent PD risk signals across 78 genomic regions^2, 3^. However, limited PD heritability is explained by these GWAS risk genes and the underlying biological mechanisms are still largely unknown. A recently developed transcriptome-wide association study (TWAS) and proteome-wide association study (PWAS) approach integrates other molecular quantitative traits such as gene expression or protein abundance with GWAS summary data, with the goal of detecting risk genes whose causal genetic effects are mediated through the considered molecular traits^4-7^.

TWAS/PWAS tools typically train genetic prediction models in reference panels containing paired genotype and molecular quantitative trait data and then integrate the resulting molecular quantitative trait loci (xQTL) weights with GWAS summary statistics to perform gene-based association tests. Particularly, standard two-sample TWAS and Mendelian randomization methods can be represented by a shared model, and these two types of analyses are shown equivalent when there is no horizontal pleiotropy effect between the molecular trait and the phenotype of interest^8^.

Recent TWASs of PD^9-12^ have nominated dozens of susceptibility genes, yet these studies relied only on bulk RNA-sequencing (RNA-seq) reference panels and often used a single statistical model to estimate expression quantitative trait loci (eQTL) weights. These features limit the feasibility and power to characterize cell-type-aware TWAS and PWAS risk genes of PD. Addressing these limitations is important because single-cell and single-nucleus studies have implicated several brain cell types - including astrocyte, microglia, and oligodendrocyte cells in the PD pathogenesis^13-15^. For example, PD-associated variants have shown cell-type-specific regulatory enrichment in mouse, including signals in microglia and oligodendroglia^14^. Single-nucleus RNA-seq studies of midbrain and cortex have identified cell-type-dependent transcriptional dysregulation, particularly in microglia and astrocytes, and enrichment of PD heritability in microglial open-chromatin regions^13, 16^. Analysis of cis-regulatory elements in the substantia nigra has additionally identified disease-perturbed pathways and highlighted potential roles for astrocytes and microglia^15^. Together, these findings motivate the integration of large bulk RNA-seq, single-nucleus RNA-seq, and proteomic reference datasets with PD GWAS summary statistics of large sample sizes.

Although degeneration of dopaminergic neurons in the substantia nigra is a defining feature of PD, the disease is increasingly recognized as a disorder of distributed brain networks extending beyond the nigrostriatal system^17^. Within these networks, altered DLPFC function and connectivity have been associated with executive and other cognitive impairments in PD, especially in later stages^18^. Thus, we will integrate the available large-scale reference omics data of DLPFC with PD GWAS summary data to enhance gene-mapping of PD. The reference omics data of DLPFC were profiled from donors in the Religious Orders Study and Rush Memory and Aging Project (ROS/MAP), including whole genome sequencing (WGS) data from whole blood, bulk RNA-seq, single-nucleus RNA-seq, and bulk proteomic data generated from DLPFC tissues^19, 20^. The use of DLPFC reference data in the present study therefore reflects both the involvement of broader cortical networks in PD and the availability of well-powered, multi-omics molecular reference panels.

We integrated the eQTL weights trained using bulk RNA-sequencing data of DLPFC from 931 postmortem brains and single-nucleus RNA-sequencing data of DLPFC from 415 postmortem brains^19, 21-23^ with PD GWAS summary statistics of European-ancestry^2^ (n=∼1.45M), to map TWAS risk genes of PD by the established omnibus TWAS (TWAS-O, see details in Methods) pipeline^24, 25^. Cell-type-aware TWAS-O was conducted using eQTL weights trained separately in six major cortical cell types: astrocytes, microglia, excitatory neurons, inhibitory neurons, oligodendrocytes, and oligodendrocyte precursor cells (OPCs). We additionally integrated protein quantitative trait loci (pQTL) weights trained using a large cohort of tandem mass tag (TMT) proteomics data (n=716) of DLPFC^25^ with the same PD GWAS summary statistics^2^ to map PWAS risk genes of PD by the analogous omnibus PWAS pipeline^26^. We next fine-mapped independent TWAS/PWAS signals by using the Gene-based Integrative Fine-mapping through conditional TWAS^27^ (GIFT) tool (see details in Methods), and examined the percentage of independent TWAS-O risk genes that have PWAS-O locus support.

To show the importance of mapping PD risk genes using reference multi-omics data of DLPFC, we performed omnibus TWAS of PD using reference bulk RNA-seq of substantia nigra (where PD’s defining pathology occurs), along with WGS genotype data from whole blood, of 183 donors from the Genotype-Tissue Expression (GTEx) V10 project^28^. We further evaluated internal support of these TWAS/PWAS risk genes of PD, using the individual-level data of genetics, global parkinsonian score (n=1,187), and a PD-related neuropathologic phenotype defined by the presence of either Lewy body pathology or nigral neuronal loss (n=1,194), from the ROS/MAP cohort. Finally, we used STRING^29^ to characterize protein-protein interaction networks and pathDIP^30^ to examine enriched biological pathways among the associated genes in DLPFC.

## Results

### Overview of the methods

We utilized our previously developed xWAS-O framework^26^ to integrate PD GWAS summary statistics from approximately 1.4 million controls^2^ with cis-xQTL weights^24, 25^ that have been trained from the ROS/MAP DLPFC multi-omics reference panels in our previous studies^19, 21-23^. The multi-omics reference panels comprised bulk RNA-seq data from (n=931), snRNA-seq data (n=415), and TMT proteomics data (n=716) from DLPFC tissues of postmortem brains in ROS/MAP. For each molecular trait, three sets of cis-xQTL weights trained by complementary statistical models (TIGAR/DPR^6^, PrediXcan/EN^4^, and the FUSION/BestModel^5^) were used to derive model-specific TWAS/PWAS p-values per gene, which are combined by the aggregated Cauchy association test (ACAT) to obtain the omnibus TWAS/PWAS p-value per gene (see details in Methods).

The TWAS/PWAS association outputs included 13,003 bulk gene-expression traits, 82,194 gene-by-cell-type traits across six cell types (13,732 in astrocytes, 13,903 in excitatory neurons, 13,669 in inhibitory neurons, 13,680 in microglia, 13,677 in oligodendrocytes, and 13,533 in OPCs), and 4,723 protein-abundance traits. Using the Aggregated Cauchy Association Test (ACAT) method^31^, bulk and cell-type-aware TWAS-O significance was defined using the genome-wide significance threshold *p*_*ACAT*_ ≤ 2.5 × 10^−6^. For PWAS-O, ACAT p-values were adjusted by genomic control factor adjustment^32^, followed by Benjamini-Hochberg procedure^33^, and *q*_*FDR*_ ≤ 0.05 defined significance since only 4,723 protein coding genes were tested by PWAS-O.

GIFT^27^ was subsequently used for locus-level conditional prioritization among multiple nearby significant genes. Within multi-gene candidate regions, genes with *p*_*GIFT*_ ≤ 0.05 were classified as independent signals. A significant gene outside the selected multi-gene GIFT candidate regions was retained as an independent single-gene candidate locus. Briefly, within each genomic region defined as merged overlapped cis-regions of nearby significant genes, GIFT jointly modeled the genetically regulated molecular traits of all test genes in the region while accounting for linkage disequilibrium (LD) in the test genome region and correlations among the considered molecular traits (see details in Methods).

We conducted exact-gene and locus-level comparisons in the study. Exact-gene overlap required an identical gene symbol, while locus-level overlap means the expanded-cis-windows (±1 Mb around gene region) of two genes overlapped on the same chromosome. Overlapped expanded-cis-window of two genes means they might share cis-regulatory eQTL in LD. Cell-type specificity was evaluated among the independent significant gene loci across the six cell types. An independent gene locus was considered cell-type-specific if its expanded-cis-window did not overlap the expanded-cis-window of any independent gene locus in the other five cell types.

### Bulk DLPFC TWAS-O findings

The quantile-quantile plots for the three model-specific bulk TWAS analyses and the omnibus analysis are shown in **Fig. S1**. Deviations from the null distribution were observed, likely due to the high polygenicity in the PD GWAS data, overlapping cis-regions of nearby genes, LD, and correlation among genetically regulated expression traits. We detected 39 marginally significant bulk TWAS-O genes (**Fig. 1A; Supplementary Data 1**), from which we fine-mapped 19 independent gene loci by GIFT or the single-gene-locus rule (**Table 1; Fig. S11**). Especially, 17 of the 39 significant genes were reported as PD-associated in the GWAS Catalog^34^ database (**Supplementary Data 1**).

**Figure 1.**
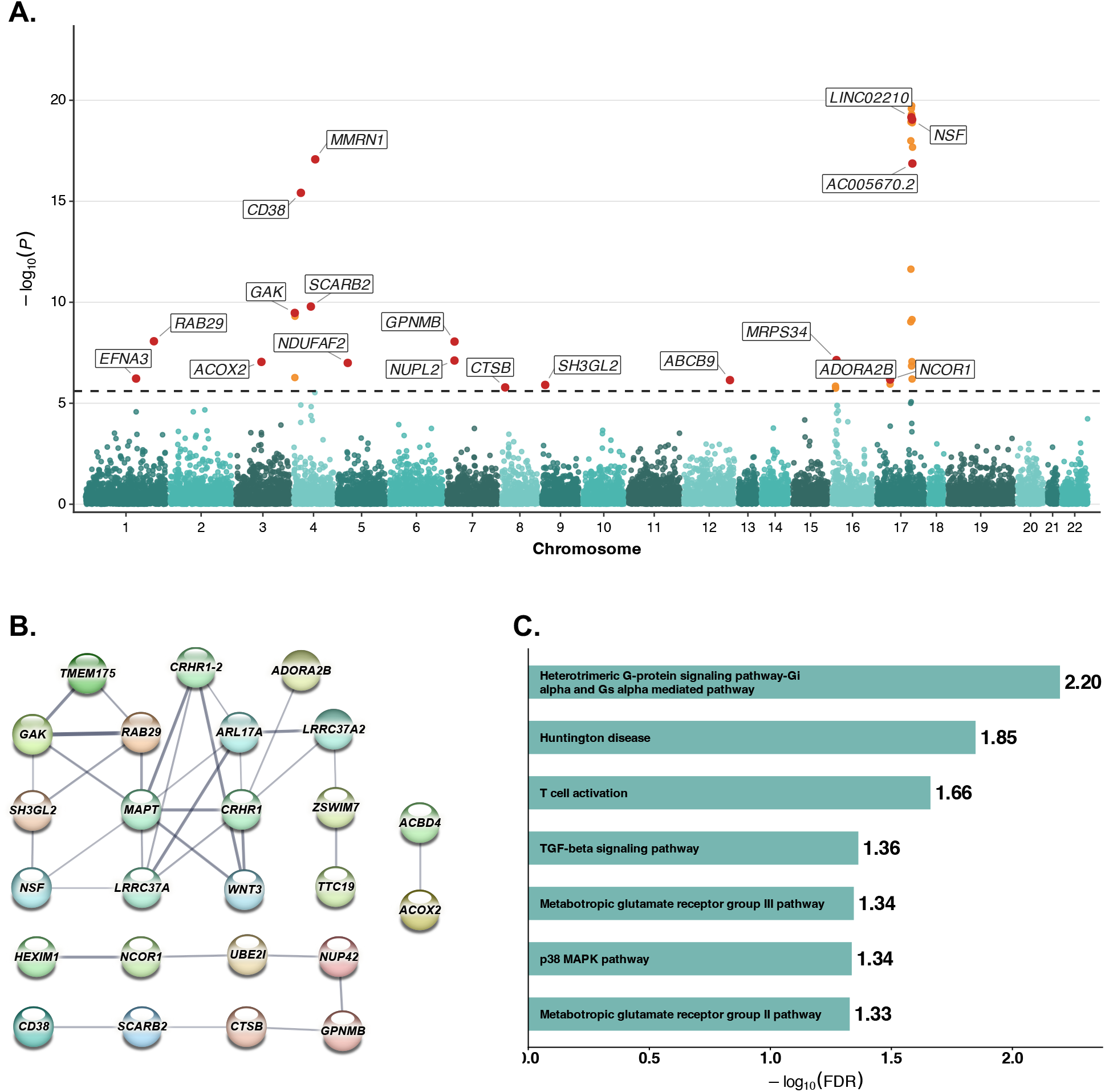
Bulk TWAS-O results in DLPFC for PD. (A) Manhattan plot of bulk TWAS-O results in DLPFC. The dashed horizontal line indicates the transcriptome-wide significant threshold of *p*_*ACAT*_ = 2.5 × 10^−6^. GIFT-prioritized or otherwise statistically independent significant genes are shown in red and labeled, whereas the remaining significant genes are shown in orange. (B) STRING functional association network constructed using the 39 significant bulk TWAS-O genes. Of these, 25 genes with at least one STRING connection are displayed. Edge thickness represents the strength of supporting evidence, quantified by the STRING confidence score. A minimum required interaction score of 0.4 was used. (C) Selected pathways significantly enriched with 39 marginal significant bulk TWAS-O genes according to PathDIP. Bars represent -log_10_(Benjamini-Hochberg FDR q-values). Complete enrichment results are provided in Supplementary Data 11.

Among the 19 independent gene loci that were colored in red and labeled in the Manhattan plot **Fig. 1A**, 7 of these (*ABCB9, AC005670*.*2, ACOX2, ADORA2B, MRPS34, NCOR1*, and *NUPL2*) were not captured by previous GWASs of PD or neurodegeneration phenotypes from GWAS Catalog but were supported by other previous PD-relevant functional studies. For example, *ACOX2* encoding a peroxisomal enzyme involved in the β-oxidation of branched-chain fatty acids^35^; *NUPL2* has relatively direct prior support by an integrative analysis of PD GWAS, brain expression, splicing, and methylation data which identified a disease-associated splicing signal at *NUPL2* and placed it in co-expression modules related to ubiquitin-dependent protein catabolism^11^; *ABCB9* was also highlighted in a recent joint-tissue integrative analysis that summarized prior evidence of PD-associated methylation^36^.

### Cell-type-aware DLPFC TWAS-O findings

Across the six cell types, we identified 133 significant gene-by-cell-type associations (21 in astrocytes, 24 in microglia, 25 in excitatory neurons, 19 in inhibitory neurons, 21 in oligodendrocytes, and 23 in OPCs, corresponding to 66 unique genes), which were fine-mapped to 43 gene-by-cell-type independent associations (10 in astrocytes, 8 in microglia, 5 in excitatory neurons, 8 in inhibitory neurons, 4 in oligodendrocytes, and 8 in OPCs, representing 32 unique genes) (**Fig. 2; Table 2; Supplementary Data 2-7**). Quantile-quantile plots are shown in **Fig. S2-S7**, and GIFT locus plots are shown in **Fig. 3** and **Fig. S12**.

**Figure 2.**
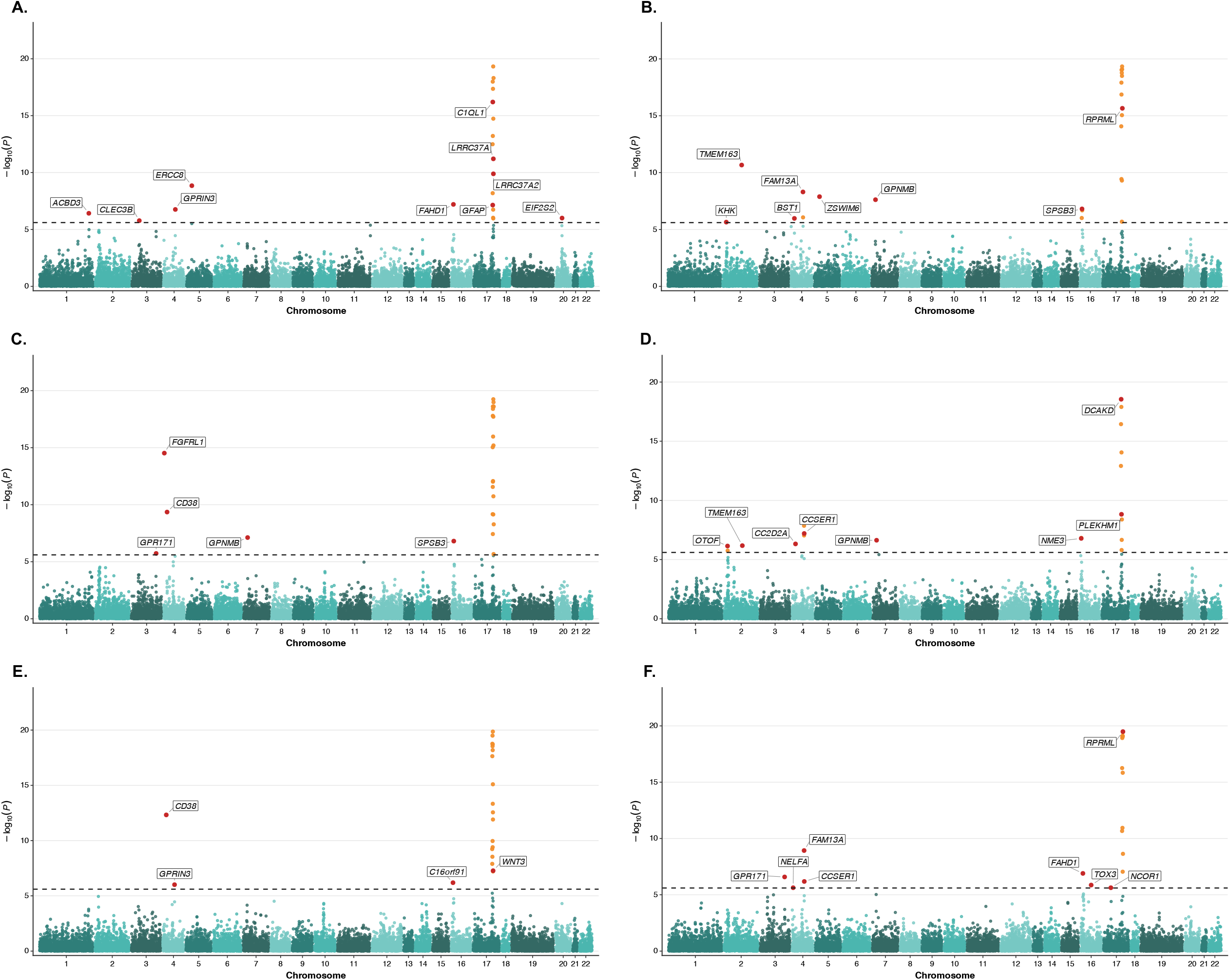
Cell-type-aware TWAS-O results in DLPFC for PD in astrocytes (A), microglia (B), excitatory neurons (C), inhibitory neurons (D), oligodendrocytes (E), and OPCs (F). Manhattan plots show cell-type-aware TWAS-O results in astrocytes, microglia, excitatory neurons, inhibitory neurons, oligodendrocytes, and oligodendrocyte precursor cells (OPCs). The dashed horizontal line indicates the transcriptome-wide significance threshold of *p*_*ACAT*_ =2.5 × 10^−6^. Significant genes prioritized as statistically independent by GIFT, together with significant genes whose expanded test regions did not overlap those of other significant genes, are shown in red and labeled. The remaining significant genes are shown in orange.

**Figure 3.**
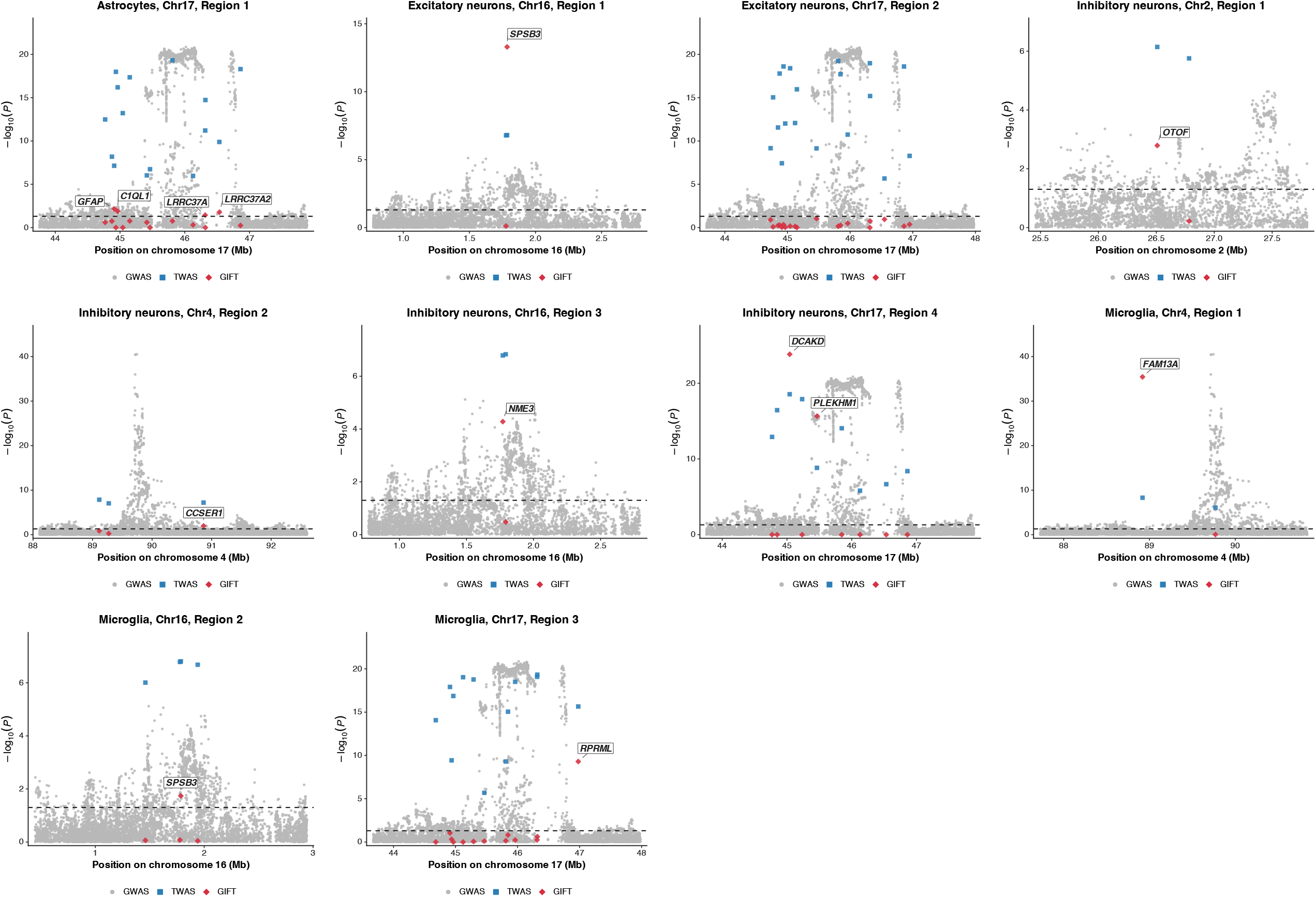
GIFT fine-mapping of cell-type-aware TWAS-O loci of PD in astrocytes, microglia, excitatory neurons, and inhibitory neurons. For each locus, GWAS SNP association results are shown as gray circles, marginal TWAS-O gene association results as blue squares, and GIFT conditional gene association results as red diamonds. The y-axis shows -log_10_(p-values), and the dashed horizontal line indicates the GIFT significance threshold of *p*_*GIFT*_ 0.05. Genes remaining significant after GIFT conditional analysis are labeled as GIFT-prioritized independent signals.

Using exact gene symbols, 13 of the 66 unique cell-type-aware genes were also significant in the bulk TWAS analysis in DLPFC: *ACBD4, ARL17A, CD38, CRHR1, GPNMB, HEXIM1, LRRC37A, LRRC37A2, MAPT, NCOR1, NSF, PLCD3*, and *WNT3*. Pairwise exact-gene Jaccard indices among the six cell types ranged from 10.3% to 36.1%, with the greatest overlaps between excitatory neurons and microglia (36.1%), astrocytes and oligodendrocytes (35.5%), and astrocytes and excitatory neurons (35.3%) (**Fig. S15**). These comparisons showed partial overlap among the significant TWAS-O gene sets across the six cortical cell types.

We assessed cell-type-specificity among the fine-mapped independent gene loci. We define an independent gene locus as cell-type-specific if its expanded-cis-window is not overlapped with the expanded-cis-window of any other independent gene loci in other cell types. We detected 3 and 2 cell-type-specific independent gene loci for astrocytes and OPCs, respectively.

We next examined independent cell-type-aware gene loci that were neither reported as PD-associated in GWAS Catalog nor supported by an expanded-cis-window overlap with a significant bulk TWAS-O gene in DLPFC. This category included *ACBD3, CLEC3B*, and *EIF2S2* in astrocytes; *KHK* in microglia; *GPR171* in excitatory neurons; *OTOF* in inhibitory neurons; and *GPR171* and *TOX3* in OPCs. Several of these findings have relevant prior biological context support. In astrocytes, *CLEC3B* encodes tetranectin, which has been reported to be reduced in the cerebrospinal fluid of patients with PD. Experimental depletion of tetranectin enhanced p53/Bax-mediated dopaminergic neuronal apoptosis and motor impairment, whereas its overexpression was neuroprotective^37^. The *EIF2S2* finding is also consistent at the pathway level with previous evidence implicating EIF2 signaling in genetically determined and sporadic PD, although that study did not specifically validate *EIF2S2* as the causal gene^38^. In microglia, *KHK*-dependent fructose metabolism has been shown to promote NOX4-mediated oxidative stress^39^, but this evidence was obtained in a model of diabetes-associated cognitive dysfunction and should be regarded as a hypothesis-generating microglial mechanism rather than PD-specific support. Among the neuronal findings, single-cell profiling of an MPTP-parkinsonism macaque model identified *OTOF*-expressing inhibitory neurons within the nigrostriatal system^40^. In OPCs, variants at the *TOX3* locus have previously been examined in a PD case-control study, with a nominal association reported for rs3104767^41^.

### Bulk PWAS-O findings in DLPFC and cross-omics support

Among 4,723 protein coding genes tested in the PWAS-O analyses, 17 genes met the genomic-control-adjusted FDR threshold, and 11 were fine-mapped as independent signals (**Fig. 4A; Table 3; Supplementary Data 8; Figs. S8 and S13**). We found that 13 of the 17 significant genes were not reported as PD-associated in GWAS Catalog, and 3 independent PWAS-O gene loci (*EXOSC7, SNX17*, and *VPS26B*) were neither reported as PD-associated in GWAS Catalog nor overlapped with a significant bulk TWAS-O finding (by both exact-gene and expanded-cis-window comparisons). Interestingly, 2 of these 3 independent PWAS-O gene loci were reported with endosomal and retromer biology relevant to neurodegeneration by previous studies: *SNX17* regulates amyloid precursor protein trafficking and processing^42^ (general neurodegenerative context support); *VPS26B*, a retromer component, has shown a protective effect against MPP+-induced toxicity in a dopaminergic cell model^43^ (PD-model-based biological plausibility support).

**Figure 4.**
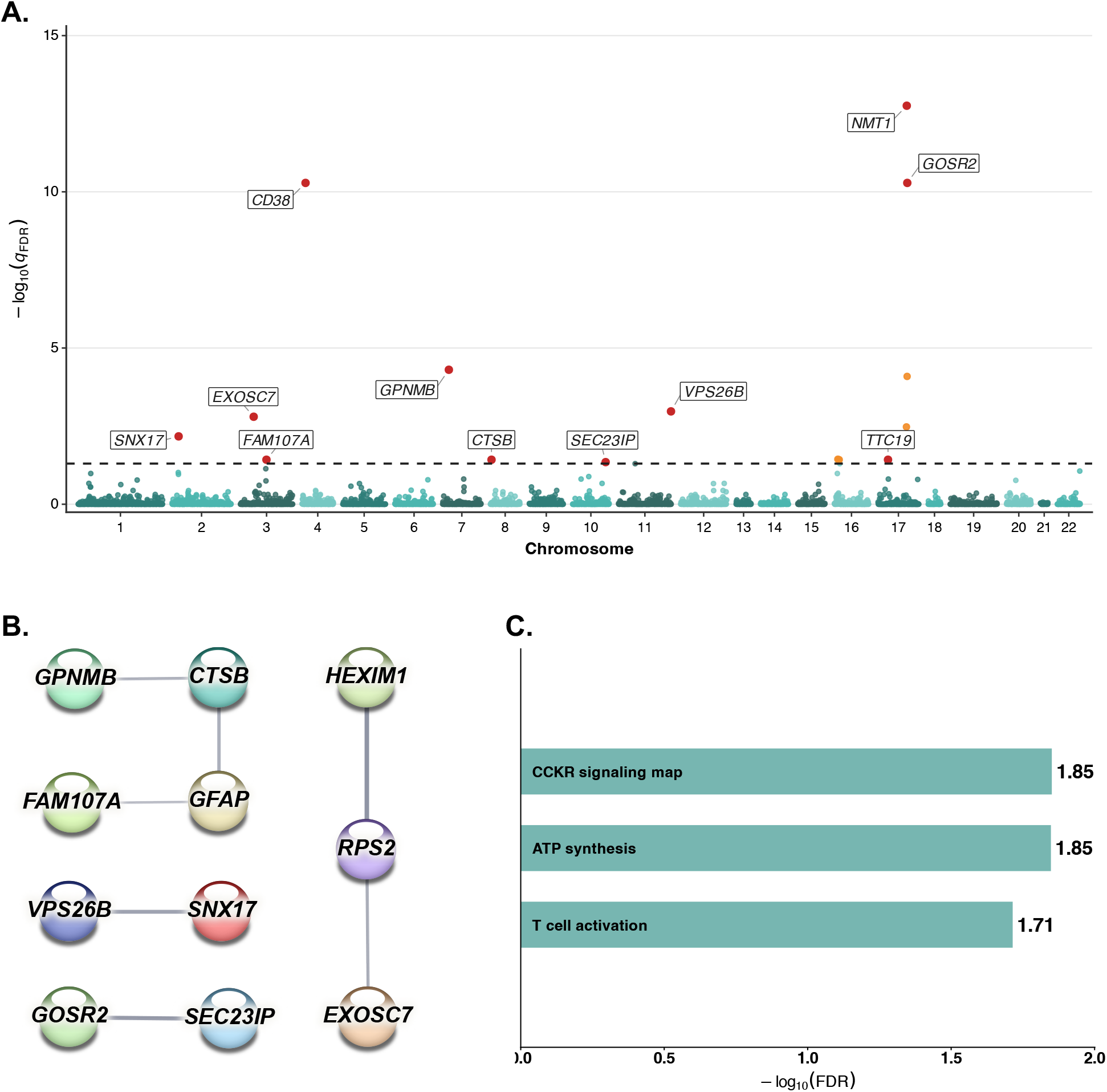
Bulk PWAS-O results in DLPFC for PD. (A) Manhattan plot of bulk PWAS-O results. The dashed horizontal line indicates the significance threshold of FDR *q* = 0.05. GIFT-prioritized or otherwise statistically independent significant genes are shown in red and labeled, whereas the remaining significant genes are shown in orange. (B) STRING functional association network constructed using the 17 significant bulk PWAS-O genes. Of these, 11 genes with at least one STRING connection are displayed. Edge thickness represents the strength of supporting evidence, quantified by the STRING confidence score. A minimum required interaction score of 0.4 was used. (C) Pathways significantly enriched with the 17 bulk PWAS-O genes according to pathDIP. Bars represent -log_10_(Benjamini-Hochberg FDR q-values). Complete enrichment results are provided in Supplementary Data 11.

By exact-gene overlap comparisons, 5 of the 17 PWAS-O genes (29.4%; *CD38, CTSB, GPNMB, HEXIM1*, and *TTC19*) were also significant in the bulk TWAS-O analysis, and six (35.3%; *CD38, GFAP, GOSR2, GPNMB, HEXIM1*, and *NMT1*) were significant in at least one cell-type-aware TWAS-O analysis (**Fig. S14**). Calculated from the combined gene sets shown in **Fig. S14**, the exact-gene Jaccard indices were 9.8% between bulk TWAS-O and PWAS-O, 7.8% between cell-type-aware TWAS-O and PWAS-O, and 14.1% between bulk and cell-type-aware TWAS-O.

By expanded-cis-window overlap comparisons, 11 of 19 (57.9%) independent bulk TWAS-O genes and 20 of 32 (62.5%) independent unique cell-type-aware TWAS-O genes have overlapped expanded-cis-window(s) with at least one of the significant PWAS-O gene, which are considered as supported by PWAS. Specifically, these 20 independent unique cell-type-aware TWAS-O genes include 6 of 10 astrocyte genes (60.0%), 5 of 8 microglial genes (62.5%), 3 of 5 excitatory-neuron genes (60.0%), 6 of 8 inhibitory-neuron genes (75.0%), 3 of 4 oligodendrocyte genes (75.0%), and 3 of 8 OPC genes (37.5%) (**Table 4**).

Among the PWAS-supported independent cell-type-aware TWAS-O gene loci, two met the cell-type-specificity criterion: *CLEC3B* in astrocytes and *NCOR1* in OPCs. Previous studies also provided biological support for other PWAS-supported cell-type-aware TWAS-O findings. For example, plasma *GFAP* has been associated with motor subtype and prognosis in early PD^44^; a *BST1* risk variant has been associated with longitudinal PD progression^45^; and *PLEKHM1* participates in autophagosome-lysosome fusion, a pathway implicated by inherited PD-associated regulatory variation^46, 47^.

### COMPARISON to bulk TWAS-O risk genes in substantia nigra

To assess cross-reference-panel support in the PD directly related substantia nigra tissue, we applied the xWAS-O framework^26^ to map PD risk genes using the reference bulk RNA-seq data from substantia nigra tissues and accompanied WGS genotype data of 183 samples from the GTEx V10 project^28^. The GTEx participants are not overlapped with the ROS/MAP cohort. Among 23,850 genes whose expression traits in substantia nigra were tested, 74 genes were detected significant with *p*_*ACAT*_ ≤2.5 × 10 ^−6^. Using the GIFT tool, we fine-mapped 27 independent gene loci, including 23 genes with *p*_*GIFT*_ ≤0.05 and four genes in single-gene candidate loci (**Fig. S9 and S10; Supplementary Data 9**).

Next, we did exact-gene comparison between the TWAS-O risk genes in substantia nigra and our findings in DLPFC. We found that these 74 significant genes in substantia nigra included 21 of 39 (53.8%) significant bulk TWAS-O genes, 24 of 66 (36.4%, 11 of 21 in astrocyte, 9 of 24 in microglial, 14 of 25 in excitatory-neuron, 7 of 19 in inhibitory-neuron, 13 of 21 in oligodendrocyte, and 10 of 23 in OPC) unique cell-type-aware TWAS-O genes, and 3 of 17 (17.6%) PWAS-O genes in DLPFC (**Fig. S16**). We note that overlapped genes are not mutually exclusive in six cell types because some genes were significant in more than one cell type (**Fig. S15**).

By expanded-cis-window comparison of independent gene loci between substantia nigra and DLPFC, we found that 13 of 19 (68.4%) independent bulk TWAS-O genes, 21 of 32 (65.6%) unique independently prioritized cell-type-aware TWAS-O genes, and 4 of 11 (36.4%) independent PWAS-O genes in DLPFC overlapped with independent gene loci in substantia nigra. The overlapping cell-type-aware findings included 7 of 10 (70.0%) independent genes in astrocytes, 5 of 8 (62.5%) in microglia, 3 of 5 (60.0%) in excitatory neurons, 5 of 8 (62.5%) in inhibitory neurons, 3 of 4 (75.0%) in oligodendrocytes, and 6 of 8 (75.0%) in OPCs. Across the six cell types, these findings represented 29 of 43 (67.4%) independent gene-by-cell-type associations, corresponding to 21 of 32 (65.6%) unique genes because some genes were independently prioritized in more than one cell type.

### Internal support of mapped PD risk genes in DLPFC

To assess whether the TWAS/PWAS risk genes in DLPFC identified from the large-scale PD GWAS summary data were also associated with motor impairment and PD-related neuropathologic traits in ROS/MAP, we performed TWAS/PWAS analyses using individual-level WGS genotype data and two phenotypes in ROS/MAP cohort - the global parkinsonian score (n=1,187) and a binary PD-related neuropathologic phenotype (n=1,194)^48,49^. The global parkinsonian score is a continuous measure of motor impairment, by summarizing the evaluations of parkinsonian gait, rigidity, bradykinesia, and tremor using 26 items from a modified Unified Parkinson’s Disease Rating Scale^50, 51^. The PD-related neuropathologic phenotype was defined as the presence of either nigral neuronal loss or Lewy body pathology. All TWAS/PWAS analyses adjusted for age at death, sex, study indicator (ROS or MAP), and the first two genotype principal components.

Given the substantially smaller ROS/MAP phenotype sample sizes, we evaluated nominal internal support using *p*_*ACAT*_ ≤0.05 and concordant component-model effect directions between the ROS/MAP and primary PD GWAS-based analyses (see Methods). We found internal support for 5 unique cell-type-aware TWAS-O risk genes using the global parkinsonian scores in ROS/MAP (**Supplementary Data 10**), including TSR3 in astrocytes; *CLEC3B, CCDC43*, and *HEXIM2* in microglia; *HEXIM2* in excitatory neurons; and *NELFA* in inhibitory neurons. We also found internal support for 10 unique genes across the cell-type-aware TWAS-O and PWAS-O analyses using the PD-related neuropathologic phenotype in ROS/MAP (**Supplementary Data 10**), including 6 cell-type-aware TWAS-O associations (*ACBD3* in astrocytes, *CLEC3B* in microglia, *ACOX2* and *BST1* in excitatory neurons, and *OTOF* and *IDUA* in OPCs), and 4 proteomic PWAS-O (*CLCN7, RPS2, MLST8*, and *ADAM11*).

### PPI network and enrichment analyses

We used STRING^52^ to examine known and predicted protein-protein interactions corresponding to the significant bulk TWAS-O, PWAS-O, and unique cell-type-aware TWAS-O genes. Genes with at least one STRING connection are displayed in the network plots, with edge thickness indicating the interaction confidence score (**Figs. 1B, 4B**, and **5A**). We separately used pathDIP^30^ to identify pathways enriched with these three gene sets at Benjamini-Hochberg FDR *q*< 0.05. Selected PD related pathways enriched with bulk and cell-type-aware TWAS-O gene loci are presented in **Fig. 1C** and **Fig. 5B**, and complete enrichment results are provided in **Supplementary Data 11**. All three significant pathways enriched with PWAS-O risk genes are displayed in **Fig. 4C**.

**Figure 5.**
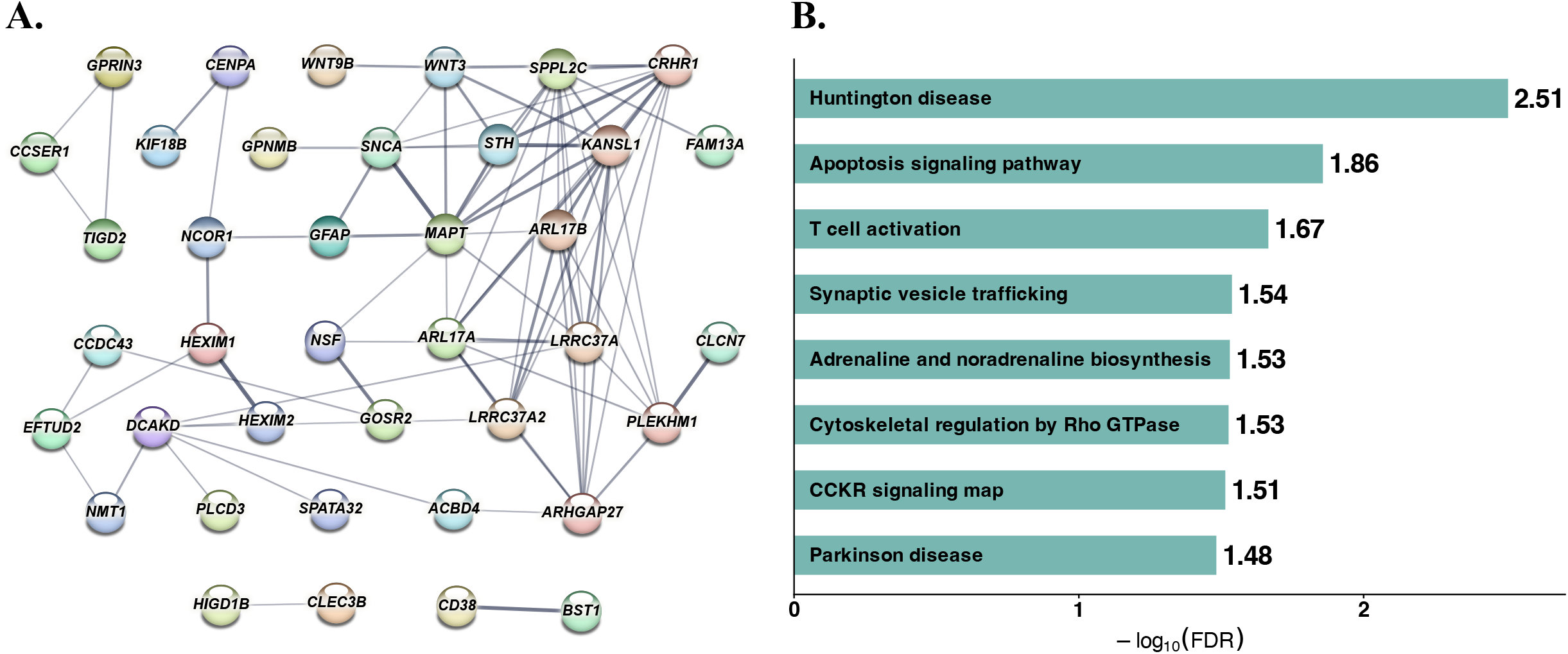
Protein-protein interaction networks and pathway enrichment results for cell-type-aware TWAS-O genes in DLPFC for PD. Significant cell-type-aware TWAS-O genes from astrocytes, microglia, excitatory neurons, inhibitory neurons, oligodendrocytes, and oligodendrocyte precursor cells (OPCs) were combined, resulting in 66 unique genes. (A) STRING functional association network constructed using the 66 unique cell-type-aware TWAS-O genes. Of these, 39 genes with at least one STRING connection are displayed. Edge thickness represents the strength of supporting evidence, quantified by the STRING confidence score. A minimum required interaction score of 0.4 was used. (B) Selected pathways significantly enriched with the full set of 66 unique cell-type-aware TWAS-O genes according to pathDIP. Bars represent -log_10_(Benjamini-Hochberg FDR q-values). Complete enrichment results are provided in Supplementary Data 11.

Among the 39 bulk TWAS-O genes, 25 (64.1%) had at least one STRING connection (**Fig. 1B**). The network contained a large component involving *MAPT, CRHR1, NSF, LRRC37A, LRRC37A2, WNT3, GAK, RAB29, TMEM175*, and *SH3GL2*, together with several smaller connected modules. PathDIP identified 10 significant pathways for the bulk TWAS-O gene set (**Supplementary Data 11**). The most significant was *heterotrimeric G-protein signaling through Gi/Gs alpha subunits* (FDR=6.36E-03; with leading risk genes of *ADORA2B, CRHR1, GAK*, and *SH3GL2*), which is relevant to striatal cAMP regulation and motor loop imbalance in PD^53^. Additional pathways shown in **Fig. 1C** included *Huntington disease* (FDR=1.42E-02), *T-cell activation* (FDR=2.18E-02), *TGF-beta signaling* (FDR=4.33E-02), *metabotropic glutamate receptor group III signaling* (FDR=4.52E-02), *p38 MAPK signaling* (FDR=4.61E-02), and *metabotropic glutamate receptor group II signaling* (FDR=4.70E-02).

Among the 17 PWAS-O genes, 11 (64.7%) had at least one STRING connection and formed 4 smaller modules rather than a single dense network (**Fig. 4B**): *GPNMB*-*CTSB*-*GFAP*-*FAM107A, HEXIM1*-*RPS2*-*EXOSC7, GOSR2*-*SEC23IP*, and *VPS26B*-*SNX17*. PWAS-O pathways include *CCKR signaling map* (FDR=1.41E-02; *CD38, GFAP, RPS2*, and *SNX17*), *ATP synthesis* (FDR=1.42E-02; *HEXIM1* and *PDHB*), and T-cell activation (FDR=1.93E-02; *CD38, PDHB*, and *SNX17*), as shown in **Fig. 4C**. These findings highlight signaling, immune, and mitochondrial-energy-related annotations within the smaller proteomic gene set.

Among the 66 unique cell-type-aware TWAS-O genes, 39 (59.1%) had at least one STRING connection (**Fig. 5A**). Most connected genes formed a broad network containing established PD-associated genes such as *MAPT*^54^, *SNCA*, and *LRRC37A2*^55^ together with genes detected across neuronal and glial panels, including *GFAP, CD38, BST1, NSF, PLEKHM1*, and *GPNMB*. PathDIP identified 14 significant pathways for this gene set (**Supplementary Data 11**). Selected pathways displayed in **Fig. 5B** included *Huntington disease* (FDR=3.11E-03), *apoptosis signaling* (FDR=1.39E-02), *T-cell activation* (FDR=2.16E-02), *synaptic vesicle trafficking* (FDR=2.90E-02), *adrenaline and noradrenaline biosynthesis* (FDR=2.95E-02), *cytoskeletal regulation by Rho GTPase* (FDR=2.98E-02), the *CCKR signaling map* (FDR=3.06E-02), and the *Parkinson disease* pathway (FDR=3.29E-02). The synaptic-vesicle and catecholamine-related enrichments were driven by *NSF* and *SNCA*, whereas *MAPT, SNCA, MAP3K14*, and other genes contributed to several of the broader disease, immune, and signaling annotations.

Comparison across molecular layers revealed both shared and omics-specific enrichment patterns. *T-cell activation*^56^ was significantly enriched with all three sets of risk genes, *the CCKR signaling map*^57^ was enriched with the PWAS-O and cell-type-aware TWAS-O gene sets, and *Huntington disease*^58^ was enriched with bulk and cell-type-aware TWAS-O gene sets. Together, these overlaps point to recurring immune, receptor-signaling, neurotransmitter/vesicle, and energy-metabolism themes as revealed by transcriptomics of bulk and single nucleus and bulk proteomics data of DLPFC, and GWAS summary data of PD.

## Discussion

### Enhanced gene-mapping of PD by integrative omics analyses

We integrated large-cohort bulk RNA-seq^21^ (n=931), snRNA-seq^19^ (n=415), and bulk TMT proteomics data^20^ (n=716) of DLPFC with the latest GWAS summary data of PD^2^ through the xWAS-O framework^26^, with the goal of enhancing the gene-mapping of PD. The reference omics data of DLPFC contained substantial participant overlap: 405 of the 415 snRNA-seq participants were included in the bulk RNA-seq panel, 677 of the 716 proteomics participants were included in the bulk RNA-seq panel, and 327 participants contributed to both the snRNA-seq and proteomics panels (**Supplementary Data 12**). Risk genes detected from these integrative omics analyses have genetic effects that are potentially mediated through bulk or cell-type-aware gene expressions or protein abundances in DLPFC.

We identified 39 significant risk genes by bulk TWAS-O, 66 unique significant genes by cell-type-aware TWAS-O, and 17 significant genes by PWAS-O, which were respectively fine-mapped by GIFT or the prespecified single-gene-locus rule to 19, 32, and 11 independent gene loci (**Fig. S16**). By exact-gene comparison, 5 of the 17 PWAS-O genes were also significant in the bulk TWAS-O analysis and 6 were significant in at least one cell-type-aware TWAS-O analysis. By expanded-cis-window comparison, 11 of 19 (57.9%) independent bulk TWAS-O genes and 20 of 32 (62.5%) unique independent cell-type-aware TWAS-O genes had an expanded-cis-window overlap with at least one significant PWAS-O gene. These overlaps provide cross-omics support for the TWAS-O/PWAS-O risk genes of PD mapped by integrating transcriptomics and proteomics data with GWAS summary data of PD.

Most independent cell-type-aware TWAS-O gene loci were supported by expanded-cis-window overlap with significant bulk TWAS-O genes in DLPFC (**Table 4**). Pairwise exact-gene comparisons also showed overlap among the significant cell-type-aware TWAS-O gene sets across the six cell types (**Fig. S15**). These findings suggest that some PD-associated genetic signals may be shared across cortical cell types. A previous stratified-LDSC analysis integrating snRNA-seq study across multiple brain cell types including astrocytes, microglia, and oligodendrocytes etc.^59^ also showed that PD risk loci are not enriched in genes specific to individual brain cell types or regions, but rather in gene sets involved in global cellular processes, especially those related to lysosomal function and genes intolerant to loss-of-function mutations, which are broadly expressed across multiple glial and neuronal subtypes. This previous study also showed that PD heritability did not localize to a single brain cell type but instead enriched in global processes of lysosomal, autophagy and LoF-intolerant pathways^59^.

### Cell-type-specific TWAS-O gene loci were mapped

Nevertheless, we identified five cell-type-specific independent TWAS-O gene loci in DLPFC: *ACBD3, CLEC3B*, and *EIF2S2* in astrocytes; and *TOX3* and *NCOR1* in OPCs. These loci were independently prioritized in their respective cell-type analyses, and their expanded-cis-windows did not overlap those of any independent gene loci in the other five cell types. No independent loci met this cell-type-specificity criterion in microglia, excitatory neurons, inhibitory neurons, or oligodendrocytes. Here, cell-type specificity describes the pattern of independent associations across the six considered cell types and does not establish that the corresponding genes have biological effects exclusively in these cell types.

### Interpretation of signals at chromosome 17q21.31

A notable concentration of significant and independently prioritized signals occurred within or near chromosome 17q21.31. The detected independent gene loci of DLPFC in this region included *LINC02210* and *NSF* in bulk tissue, *LRRC37A* and *LRRC37A2* in astrocytes, and *WNT3* in oligodendrocytes, while the broader significant sets also contained *MAPT, CRHR1, ARL17A*, and other neighboring genes. This region contains the approximately 900-kb MAPT inversion that defines the extended H1 and H2 haplotypes and is further characterized by segmental duplications and copy-number variation. The inversion produces distinct and extended linkage-disequilibrium patterns and can influence expression of multiple genes in a tissue-dependent manner^60^; H1 sub-haplotypes have also been associated with PD risk and with regulation of *LRRC37A/LRRC37A2* in astrocytes and human brain tissue^55^. Consequently, several gene-level associations in this region may reflect a shared haplotypic or cis-regulatory architecture rather than multiple fully separable causal mechanisms. GIFT reduces redundancy by jointly evaluating correlated genes within each tested molecular panel, but an “independent” GIFT signal does not establish independence across cell types or multiple omics layers and may not fully resolve the complex inversion. Follow-up structural-haplotype-aware fine-mapping, colocalization, and functional studies will therefore be helpful for better characterizing the contributions of individual genes at this locus.

### Relevance of the multi-omics reference panels of DLPFC

Although degeneration of dopaminergic neurons in the substantia nigra is a defining feature of PD, it is known that PD pathology is not confined only to the substantia nigra. The PD pathology can start in the brainstem/olfactory system and eventually involve cortical regions such as the DLPFC, rather than isolated in substantia nigra^18^. Especially, PD also affects distributed cortico-subcortical systems involved in executive, cognitive, and motor-related function^61^. Our main analyses in this work leverage the available large-scale multi-omics reference data of DLPFC in the ROS/MAP cohort^19, 20^ to enhance the gene-mapping of PD. To provide support of the gene-mapping using the multi-omics data of DLPFC, we also conducted analogous analyses using the bulk RNA-seq data (n=183) of substantia nigra tissues from the independent GTEx V10 project.

By exact-gene comparison of independently prioritized TWAS-O findings, we identified five genes (*ADORA2B, GAK, LINC02210, NDUFAF2*, and *RAB29*) that met the independence criteria in both the bulk DLPFC and GTEx substantia nigra analyses. Two additional genes independently prioritized in the cell-type-aware DLPFC analyses, *C1QL1* in astrocytes and *WNT3* in oligodendrocytes, also met the independence criteria. Exact-gene comparisons between all significant GTEx TWAS-O genes and the significant genes from each DLPFC cell-type-aware analysis showed the highest Jaccard similarities for excitatory neurons (16%) and oligodendrocytes (16%). These concordances provide support for the PD risk genes mapped from considering the omics data in DLPFC.

### Internal support in ROS/MAP

The internal TWAS/PWAS analyses using individual-level genotype and phenotype data of parkinsonian score and PD-related neuropathologic phenotype in ROS/MAP provided internal support for 5 unique genes in relation to the global parkinsonian score and 10 unique genes in relation to the PD-related neuropathologic phenotype. These genes have nominal significant TWAS/PWAS p-values < 0.05 and concordant effect size directions as the ones derived from public GWAS summary data of PD. Especially, *CLEC3B* showed internal support for both phenotypes, whereas *HEXIM2* showed internal support for the global parkinsonian score in microglia and excitatory neurons. The power is limited in these internal analyses due to small sample sizes (∼1K). These findings remain exploratory because the nominal TWAS/PWAS P-values were not corrected for multiple testing.

### PPI and pathway analyses illustrate biological functions of TWAS-O/PWAS-O signals

Further PPI network analyses of our bulk TWAS-O, cell-type-aware TWAS-O, and PWAS-O risk genes detected inter-connected PPI networks with hub genes of known PD risk genes, such as *MAPT*^*54*^, *SNCA*^*62*^, and *LRRC37A*^*55*^. Interestingly, gene pathway enrichment analyses identified several PD-related pathways that were significantly enriched with xWAS-O findings based on multiple reference omics data - *T-cell activation, CCKR signaling map*, and *Huntington Disease*. Collectively, these findings emphasize both shared and complementary insights from different omics layers in shaping PD risk, through common gene pathways.

### Limitations of this study

Despite the strength of this study, it has several important limitations common to all TWAS and PWAS studies that rely on GWAS summary statistics. We retained the prespecified stringent threshold of *P*_*ACAT*_ ≤ 2.5 × 10 ^− 6^ for the bulk and cell-type-aware TWAS-O analyses and used genomic-control-adjusted Benjamini-Hochberg FDR for PWAS-O. Inflated marginally significant gene sets arise from strong polygenic PD associations, linkage disequilibrium, overlapping cis-regions, and correlated genetically regulated molecular traits. GIFT was then used to prioritize conditionally independent signals within associated regions. Importantly, GIFT addresses correlation among nearby predicted molecular traits during locus-level prioritization, but it is not a correction for global test-statistic inflation.

Second, the reference transcriptomics and proteomics data were profiled from a single prefrontal cortical region, and it would be important to profile more than one site including other sites within and outside the brain given the widespread sites that accumulate PD pathology. Our study did not account for risk genes that act primarily in other PD-relevant brain tissues such as substantia nigra dopamine neurons or peripheral tissues. Consequently, it is unclear to what extent the phenotypic heterogeneity of PD depends on differences in sites specific underlying genetics differences.

However, our study demonstrated the feasibility of integrating multi-omics data with GWAS summary data to enhance the gene mapping for PD. The xWAS-O procedure can be easily applied to integrate omics data profiled from other brain regions and GWAS data to further enhance the gene mapping of PD and other complex diseases. Finally, functional follow-up in cellular and in-vivo systems will be essential to reveal the underlying biological mechanisms of detected risk genes.

### Conclusions

In summary, we integrated large-scale bulk RNA-seq, snRNA-seq and TMT proteomic data from the DLPFC with large-scale public PD GWAS summary data^2^ using the xWAS-O framework. We identified 39 significant bulk TWAS-O genes, 66 unique significant cell-type-aware TWAS-O genes, and 17 significant PWAS-O genes, from which 19, 32, and 11 independent candidate signals, respectively, were prioritized by GIFT or the prespecified single-gene-locus rule. Cross-omics comparisons showed that 11 of 19 independent bulk TWAS-O genes and 20 of 32 independent cell-type-aware TWAS-O genes received PWAS-O locus support, indicating convergence between the transcriptomic and proteomic analyses. A subset of these findings in DLPFC also received external support from the TWAS risk genes mapped by considering the bulk RNA-seq data of substantia nigra tissue from the GTEx v10 project, as well as internal support from the TWAS/PWAS analyses using individual-level genotype and PD-related phenotype data in ROS/MAP. Together with the observed PPI networks and pathway enrichment results, these findings provide an enhanced set of PD-associated genes and loci for future investigation. Independent association studies and functional experiments will be needed to further pinpoint the contributing genes within complex loci and determine their biological roles in PD.

## Methods

### xWAS-O analytical framework

The xWAS-O pipeline recently developed by our group^25, 26^ can integrate multi-omics data (i.e., bulk-RNAseq, sn-RNAseq, and bulk proteomics) with GWAS summary data using multiple complementary statistical methods. In brief, Stage I of xWAS-O trains cis-xQTL imputation models using individual-level genotype and transcriptomic/proteomic expression data. Stage II performs gene-based association testing by integrating estimated xQTL weights with GWAS summary statistics data, followed by omnibus p-values aggregation through ACAT^31^. This framework has been shown to improve statistical power and maintain well-calibrated type I error rates^26^.

#### Stage I. xQTL weight estimation

As described in our previous studies^24^, for each gene and for each corresponding molecular trait, cis-xQTL weights were estimated using genotype data (G) within ±1 Mb of the transcription start/termination site as predictors.

Each molecular trait was normalized and log2-transformed as appropriate for its assay platform, after which prespecified biological and technical covariates were regressed out (e.g., age, sex, batch effects, PEER factors^63^ and genotype PCs). The resulting residuals were used as the quantitative traits, ***T***_***g***_, for cis-xQTL model training. Three complementary statistical methods were implemented to estimate cis-xQTL weights (***Equation 1***): (i) Nonparametric DPR implemented in TIGAR^6,7^, (ii) Penalized regression with EN penalty implemented in PrediXcan^4^; (iii) the best predictive model selected from FUSION, which considers penalized regression with LASSO, EN, BLUP, and Top1-QTL^5^.

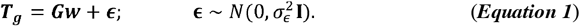

#### Stage II. Gene-based Association testing

Genes with CV *R*^2^ >0.5% for their corresponding molecular traits were retained for association testing. As shown by the TIGAR-V2 paper^7^, a relaxed threshold does not inflate type I error, because TWAS Z-scores are linear combinations of GWAS statistics weighted by estimated eQTL effects, imprecise weights reduce power but do not introduce bias.

In Stage II, xWAS-O first uses these three sets of xQTL weights 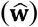 in ***Equation 1*** from Stage I to predict the genetically regulated genetic component of expression (GReX) in the GWAS test data by *G*_*gwas*_*w*, and then test the association between the phenotype *Y* and the GReX component as follows:

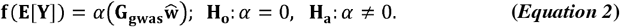

When individual genotype data of the GWAS cohort (*G*_*gWas*_) are not available, it is equivalent to take 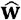 as variant weights to implement gene-based burden test as shown in the S-PrediXcan paper^64^. A gene-based association Z-score test statistic can be calculated using the summary-level GWAS test data as follows:

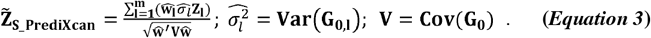

Here, **Z**_1_ denotes the single-variant Z-score test statistics in GWAS summary data for the *l*^*th*^ genetic variant, 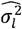 denotes genotype variance of the *l*^*th*^ genetic variant, and **v** denotes the genotype variance-covariance matrix. Both 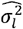 and **v** could be approximated from individual-level genotype data of an external reference panel of the same ancestry as the test GWAS data (with genotype matrix **G**_**0**_).

Reference LD derived from the WGS genotype data of the ROS/MAP cohorts^21^ was used for deriving xWAS Z-score test statistics in this study. Gene-based association test (two-sided) p-values can be easily derived from the Z-score statistic in ***Equation 3*** that has a standard normal distribution under the null hypothesis of no association exists between genetically regulated molecular traits and the phenotype of interest.

#### Derive omnibus xWAS p-values

As shown by our previous studies^26, 65^, there is no single statistical method that can achieve optimal performance of modeling the underlying genetic architecture of genome-wide genes. Thus, to enhance the robustness and statistical power of gene-based association testing, xWAS-O utilizes the ACAT^31^ to combine p-values from all three imputation tools based on complementary statistical methods as:

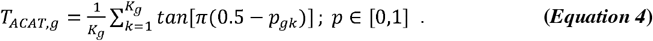

Here, *p*_*gk*_ denotes the two-sided association p-value from the *kth* contributing model for gene *g*, and *K*_*g*_ ∈ {1, 2, 3} denotes the number of available eligible xWAS p-values per gene. The ACAT test statistic (*T*_*ACAT*_) approximately follows a standard Cauchy distribution^31^ under the null hypothesis.

The omnibus test p-value for each test molecular trait of a test gene was approximated using the upper-tail probability of a standard Cauchy distribution by ***Equation 5*** and is then used to identify significant xWAS risk genes^26^. When only one model was available, the omnibus p-value equaled that model’s association p-value.

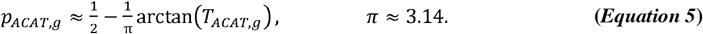

By comprehensive simulation studies and real studies of complex traits, previous studies^26, 65^ have shown that improved power and calibrated type I error rates were obtained using ACAT omnibus xWAS p-values.

### ROS/MAP reference multi-omics data and xQTL weight in DLPFC

Large-scale multi-omics reference data of DLPFC in the ROS/MAP cohort were considered in this study. The bulk transcriptomic reference panel included RNA-seq data from the DLPFC of 931 ROS/MAP participants. Gene expression was quantified as transcripts per million, log2-transformed, and adjusted for age at death, sex, postmortem interval, sequencing batch, and 20 PEER factors^24^. The cell-type-aware reference panel was derived from snRNA-seq of 436 postmortem DLPFC specimens. Raw counts were aggregated across nuclei from the same participant and cell type to generate pseudo-bulk expression traits for astrocytes, microglia, excitatory neurons, inhibitory neurons, oligodendrocytes, and oligodendrocyte precursor cells; these traits were converted to counts per million and log2-transformed. After matching to quality-controlled WGS data, 415 participants were available for weight training. Sex, age at death, study indicator (ROS or MAP), and 20 PEER factors were regressed out from each cell-type-aware expression trait^19,25^.

The proteomic reference panel included TMT mass-spectrometry data from the DLPFC of 716 ROS/MAP participants. Protein abundance was normalized relative to the sample-specific total protein abundance; proteins with measurements missing in more than 50% of participants were excluded. The abundance ratios were log2-transformed. Sex, age at death, postmortem interval, mass-spectrometry batch/quantification mode (MS2 versus MS3), and the first five genotype principal components were regressed out^20, 25^.

The cis-xQTL weights have been generated (Stage I) from these multi-omics reference data in our published xWAS studies of Alzheimer’s disease^24^. The xQTL weight estimation only involves the genotype and molecular quantitative trait data. Quality-controlled ROS/MAP WGS data were used for cis-xQTL model training in our previous studies of Alzheimer’s disease. Variants with minor-allele frequency greater than 1% and Hardy–Weinberg equilibrium *P* ≥ 1 × 10 ^− 5^ were retained. For every bulk expression, cell-type-aware expression, or protein-abundance trait, variants within 1 Mb upstream of the gene start through 1 Mb downstream of the gene end were considered cis predictors. We trained three complementary prediction models: TIGAR/DPR; elastic-net regression with equal L1 and L2 penalties; and FUSION/BestModel, selected according to five-fold cross-validation *R*^2^ from Top1-QTL, LASSO, elastic-net, and BLUP candidate models. BSLMM was not included.

The present study integrated these estimated xQTL weights given by models with five-fold cross-validation *R*^2^ > 0.5%, with the PD GWAS data to map risk genes of PD (Stage II). For each tested molecular trait, our xWAS-O method^26^ combined the association p-values obtained from these three prediction models, reducing dependence on any single model and improving power relative to model-specific analyses^26^.

### Fine-map xWAS-O risk genes by GIFT

To fine-map risk genes within genomic regions containing multiple significant xWAS-O signals, we applied Gene-based Integrative Fine-mapping through conditional TWAS, referred to as GIFT^27^. First, for each candidate locus containing multiple significant xWAS-O risk genes, we considered the merged test regions of all nearby significant xWAS-O risk genes whose test regions were overlapped with the top significant risk gene. GIFT then jointly models the genetically regulated expression or protein abundance of all significant xWAS-O risk genes in the region, explicitly accounting for both LD structure and correlations among predicted molecular traits. As a result, GIFT can effectively pinpoint conditionally independent signals. Within a multigene candidate locus, a gene with *P*_*GIFT*_ ≤ 0.05 was considered a conditionally independent signal. A significant xWAS-O gene that did not enter a multigene GIFT candidate region because its expanded test interval did not overlap that of another significant gene was classified as an independent single-gene candidate locus and did not receive a GIFT p-value. GIFT was used for locus-level conditional prioritization while accounting for local LD and correlations among predicted molecular traits; it was not used as a genomic-control correction for global test-statistic inflation.

Here, we briefly describe the underlying model assumptions by GIFT for using summary xQTL and GWAS data. For each gene *i*, the marginal association Z-scores from the xQTL data can be modeled as:

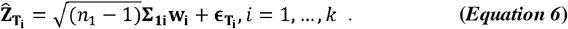

Here, *n*_*1*_ is the sample size for xQTL summary data, **∑**_**1*i***_ is the LD correlation matrix of cis-SNPs for gene *i*, **w**_**i**_ represents the vector of cis-SNP effect sizes on gene expression or protein abundance as in ***Equation 1***, and 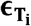 captures residual errors.

The marginal Z-score statistics of all cis-SNPs in the target genetic region from the GWAS summary data can be modeled as:

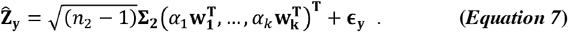

Here, *n*_2_ is the sample size for GWAS summary data; **∑**_**2**_ is the LD correlation matrix of all cis-SNPs; **w**_**i**_ are xQTL effect sizes defined in ***Equation 6***; *α*_*i*_ denotes the conditional effect of the GReX or genetically regulated protein abundance (GRP) of gene *i* on the phenotype, while accounting for the genetic components of all neighboring genes (*α* _−*i*_) in the same multiple linear regression model; and **ϵ**_**y**_ represents residual errors. A gene with significant conditional effect a_i_ will be the independent signal prioritized by GIFT. A gene with most significant marginal TWAS-O p-value may not always be the one prioritized by GIFT in a joint model of all significant genes in the same target genetic region as in ***Equation 7***.

Since our omnibus framework does not provide a combined xQTL weights, we implemented GIFT with xQTL and GWAS summary data. We generated xQTL summary data 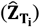 and the corresponding LD (**∑**_**1*i***_) from the individual-level snRNA-seq, TMT proteomics, and whole genome sequencing data of ROS/MAP samples^21^. We used GWAS LD (**∑**_**2**_) derived from the UK Biobank^66^ reference panel. Reference LD derived from different reference panels are recommended by GIFT developers for resolving potential computational issues.

After independent signals were identified within each cell-type panel, we assessed cell-type specificity by comparing their expanded-cis-windows across the six cell types. For each gene, the expanded-cis-window extended from 1 Mb upstream of the gene start to 1 Mb downstream of the gene end. Overlap comparisons were performed between windows on the same chromosome. An independent gene locus was classified as cell-type-specific if its expanded-cis-window did not overlap that of any independent gene locus in the other five cell types.

### PPI network analyses

Protein-protein interaction (PPI) networks were constructed for significant risk genes using the STRING^29, 52, 67^ webtool (version 12.0), which integrates multiple sources of functional association evidence, including curated databases, experimental data, text mining, co-expression, gene neighborhood or co-occurrence, and homology, to derive an overall confidence score for each protein–protein association. STRING edges therefore represent evidence that two proteins may contribute to a shared biological function and do not necessarily indicate direct physical binding. We performed STRING analyses using the complete lists of significant bulk TWAS-O genes, unique cell-type-aware TWAS-O genes across the six brain cell types, and bulk PWAS-O genes. The minimum required interaction score was set to 0.4. Networks were displayed using the confidence view, in which edge thickness represents the STRING confidence score. Genes without reported connections were omitted from the network visualizations but remained included in the corresponding pathway enrichment analyses.

### Enrichment analyses by pathDIP

PathDIP^68^ integrates pathways from 6,500□plus curated records across many databases, unifying them with a KEGG□/Reactome□based ontology so enrichment results can be filtered, consolidated, and mapped into 53 functional categories, with FDR q-values calculated by the Benjamini-Hochberg method. Specific pathway sources can be selected for gene enrichment analyses which aim to detect pathways that are significantly enriched with the curated gene list versus a similar list of random genes. We utilized the pathDIP webtool to conduct gene pathway enrichment analyses with respect to databases of Panther^69^, PathBank^70^, and Reactome^71^ with the lists of bulk TWAS-O, cell-type-aware TWAS-O, and PWAS-O risk genes of PD. For visualization, selected significant and biologically relevant pathways were plotted as -log_10_(FDR q-value); complete enrichment results are reported in **Supplementary Data 11**.

### Independent reference bulk RNA-seq data substantia nigra from GTEx V10

To assess whether the PD-associated transcriptomic signals were supported in a disease-relevant brain region from an independent cohort, we analyzed bulk RNA-seq and matched genotype data from the substantia nigra of 183 donors in the Adult GTEx V10 release^28^. Expression residuals were generated after regressing out sex, age, body mass index, and the top expression principal components. And the following Stage I model training and Stage II association studies process uses the exact same TWAS-O workflow and the same PD GWAS summary data, it was interpreted as external support in PD related substantia nigra tissue.

### Internal support from ROS/MAP with respect to global parkinsonian score and PD-related neuropathologic phenotype

As described previously, the global parkinsonian score was assessed annually by trained nurse clinicians using 26 items from a modified motor section of the Unified Parkinson’s Disease Rating Scale^50, 51^. The score summarizes four parkinsonian signs: parkinsonian gait, rigidity, bradykinesia, and tremor. The binary PD-related neuropathologic phenotype used in this study was defined as the presence of either nigral neuronal loss or Lewy body pathology. This study-specific phenotype therefore captures PD-related neuropathologic features and should not be interpreted as a clinical PD diagnosis or as the more restrictive neuropathologic definition requiring both features.

A score for each of the four parkinsonian signs was based on the sum of the scores for each of its individual items assessed, e.g., 8 items for bradykinesia. The scores for the 4 parkinsonian signs were averaged to provide a continuous global parkinsonian score as previously described.^50^ The distribution of global parkinsonian score was positively skewed and was square root transformed prior to our analyses.

Nigral neuronal loss was assessed semi-quantitatively on a 0-3 scale in hematoxylin-and-eosin-stained 6-µm sections of the substantia nigra from a hemi-section of the mid-to-rostral midbrain near the exit of the third cranial nerve. Lewy body pathology was considered present if detected in any of six sampled regions: the substantia nigra, anterior cingulate cortex, entorhinal cortex, midfrontal cortex, superior or middle temporal cortex, or inferior parietal cortex, using a monoclonal antibody against phosphorylated α-synuclein^48^.

xWAS-O analyses were performed using individual-level WGS genotype data and phenotypic data of global parkinsonian score and PD-related neuropathologic phenotype from ROS/MAP. Linear regression was used for Stage II association tests of the square-root-transformed global parkinsonian score, and logistic regression was used for Stage II association tests of the binary PD-related neuropathologic phenotype. All models adjusted for age at death, sex, study indicator (ROS or MAP), and the first two genotype principal components.

Given the much smaller ROS/MAP phenotype sample sizes than the sample size of the public GWAS of PD, we used *p*_*ACAT*_ < 0.05 as a nominal threshold and concordant effect sizes for internal support. Because ACAT combines P-values without providing an effect direction, effect size concordance was assessed using the test statistics based on individual statistical models, where TWAS P-values given by individual statistical models were not required to be below 0.05. Effect size concordance was evaluated separately for each gene and molecular panel using the available DPR, EN, and FUSION statistical models. The ROS/MAP T-statistics were required to have a common sign across the available statistical models, and the PD GWAS-based Z-statistics for the same gene and molecular panel were required to have the same sign across considered statistical models.

## Author contributions

Q.L. conducted all data analyses and drafted the paper. S.T., D.A.B., N.T.S., P.L.D., V.M., A.S.B. generated the ROS/MAP omics data, and reviewed and participated in editing the final manuscript. J.Y. conceived and supervised the study, as well as edited the manuscript.

## Statements and Declarations

### Ethical considerations

The ROS/MAP studies were approved by the Institutional Review Board of Rush University Medical Center (FWA#00000482; ORA#L99032481-CR19). All individual-level ROS/MAP data analyzed in the present study were de-identified.

### Consent to participate

All ROS/MAP participants provided written informed consent and signed Anatomical Gift Act agreements and repository consents.

### Consent for publication

All authors consent for publication.

### Data availability

All omics data of ROS/MAP samples are available from Synapse (https://www.synapse.org/Synapse:syn3219045), which can be requested through www.radc.rush.edu and www.synapse.org. GWAS summary data of PD by the Nalls et al. 2019 exclude the 23andMe cohort are available from google drive (https://drive.google.com/drive/folders/10bGj6HfAXgl-JslpI9ZJIL_JIgZyktxn). Trained bulk cis-eQTL weights, cell-type-aware cis-eQTL weights, cis-pQTL weights are available from SYNAPSE (syn66644078; https://doi.org/10.7303/syn66644078). GTEx V10 resources were available from the GTEx Portal(https://gtexportal.org/home/). Omnibus TWAS/PWAS summary data of PD will be shared through SYNAPSE once this work is published.

### Code availability

Analysis scripts used to produce all results in this work is available at https://github.com/Leo-LiuQiang/CTS-TWAS.

### Declaration of generative AI and AI-assisted technologies in the manuscript preparation process

During the preparation of this work, the author(s) used ChatGPT for contents grammar check and paraphrasing. The author(s) reviewed and edited the output as needed and take full responsibility for the content of the published article.

