## Supplementary material for "Integrating dorsolateral prefrontal cortex multi-omics and GWAS summary data enhances gene-mapping of Parkinson’s disease": Table 1-4

**Table 1. Significant bulk TWAS-O risk genes of PD in DLPFC.** The table lists 39 genes with $P_{ACAT}\leq2.5\times{10}^{-6}$. GIFT p-values are reported for multigene candidate locus; significant isolated candidates were not subjected to GIFT analysis and therefore have missing GIFT p-values.

| **Gene** | **CHR** | **Start Position** | **TWAS-O P_ACAT_** | **GIFT P-value** | **Gene** | **CHR** | **Start Position** | **TWAS-O P_ACAT_** | **GIFT P-value** |
| --- | --- | --- | --- | --- | --- | --- | --- | --- | --- |
| *EFNA3^ad^* | 1 | 155078872 | 6.00E-07 | NA | *TTC19^c^* | 17 | 15999380 | 1.13E-06 | 7.97E-01 |
| *RAB29^ad^* | 1 | 205767986 | 8.53E-09 | NA | *NCOR1^cd^* | 17 | 16029157 | 6.70E-07 | 4.36E-02 |
| *ACOX2^cd^* | 3 | 58505136 | 8.99E-08 | NA | *PLCD3^c^* | 17 | 45108967 | 9.28E-10 | 1.00 |
| *GAK^ad^* | 4 | 849276 | 3.37E-10 | 4.31E-05 | *ACBD4^c^* | 17 | 45132600 | 1.01E-18 | 1.00 |
| *TMEM175^a^* | 4 | 932387 | 5.36E-07 | 1.13E-01 | *AC142472.1^c^* | 17 | 45146730 | 2.30E-12 | 1.00 |
| *IDUA* | 4 | 986997 | 4.95E-10 | 1.61E-01 | *HEXIM1^c^* | 17 | 45148502 | 1.18E-19 | 1.00 |
| *CD38^acd^* | 4 | 15778275 | 3.82E-16 | NA | *LRRC37A4P^c^* | 17 | 45506741 | 2.57E-20 | 1.00 |
| *SCARB2^ad^* | 4 | 76158733 | 1.63E-10 | NA | *LINC02210^acd^* | 17 | 45620328 | 6.94E-20 | 1.31E-05 |
| *MMRN1^ad^* | 4 | 89879532 | 8.38E-18 | NA | *CRHR1^ac^* | 17 | 45784280 | 5.38E-20 | 1.00 |
| *NDUFAF2^ad^* | 5 | 60945129 | 1.01E-07 | NA | *MAPT-AS1^ac^* | 17 | 45799390 | 1.44E-07 | 1.00 |
| *NUPL2^cd^* | 7 | 23181827 | 7.70E-08 | 8.53E-03 | *MAPT^ac^* | 17 | 45894382 | 4.94E-20 | 1.00 |
| *GPNMB^acd^* | 7 | 23235967 | 8.83E-09 | 5.46E-03 | *MAPT-IT1^c^* | 17 | 45895783 | 1.86E-20 | 1.00 |
| *CTSB^acd^* | 8 | 11842524 | 1.65E-06 | NA | *LRRC37A^c^* | 17 | 46292733 | 8.66E-08 | 5.11E-02 |
| *SH3GL2^ad^* | 9 | 17579082 | 1.24E-06 | NA | *LRRC37A2^c^* | 17 | 46511511 | 7.20E-10 | 1.00 |
| *ABCB9^d^* | 12 | 122920951 | 7.16E-07 | NA | *ARL17A^c^* | 17 | 46516702 | 6.33E-07 | 6.24E-01 |
| *UBE2I^c^* | 16 | 1308880 | 1.42E-06 | 4.47E-01 | *NSF^acd^* | 17 | 46590669 | 9.04E-20 | 3.11E-02 |
| *TSR3^c^* | 16 | 1349240 | 1.79E-06 | 4.42E-01 | *WNT3^ac^* | 17 | 46762506 | 1.29E-19 | 1.00 |
| *MRPS34^cd^* | 16 | 1771890 | 7.29E-08 | 1.26E-04 | *AC005670.2^cd^* | 17 | 46923133 | 1.35E-17 | 1.37E-02 |
| *ADORA2B^cd^* | 17 | 15944917 | 5.41E-07 | 2.28E-02 | *LRRC37A17P^c^* | 17 | 46978481 | 2.11E-18 | 1.00 |
| *ZSWIM7^c^* | 17 | 15976560 | 3.76E-07 | 2.42E-01 |  | | | | |

- a. Listed as associated with PD traits in GWAS Catalog.
- c. Supported by a significant PWAS-O gene by expanded-cis-window overlap comparison.
- d. Independent gene loci, defined by GIFT p-value $\leq0.05$ or as a significant isolated candidate not included in a multigene GIFT analysis.

**Table 2: Independent significant cell-type-aware TWAS-O risk gene loci of PD in DLPFC.** The table lists 43 independent gene-by-cell-type associations representing 32 unique genes. GIFT p-values are reported for multigene candidate locus; significant isolated candidates were not subjected to GIFT and therefore have missing GIFT p-values.

| **Gene** | **CHR** | **Cell type** | **TWAS-O P_ACAT_** | **GIFT P-value** | **Gene** | **CHR** | **Cell type** | **TWAS-O P_ACAT_** | **GIFT P-value** |
| --- | --- | --- | --- | --- | --- | --- | --- | --- | --- |
| *ACBD3^d^* | 1 | Ast | 3.81E-07 | NA | *SPSB3^bcd^* | 16 | Ex | 1.55E-07 | 5.02E-14 |
| *CLEC3B^cd^* | 3 | Ast | 1.69E-06 | NA | *OTOF^cd^* | 2 | In | 7.15E-07 | 1.61E-03 |
| *GPRIN3^abd^* | 4 | Ast | 1.75E-07 | NA | *TMEM163^ad^* | 2 | In | 6.64E-07 | NA |
| *ERCC8^bd^* | 5 | Ast | 1.43E-09 | NA | *CC2D2A^abcd^* | 4 | In | 4.75E-07 | NA |
| *FAHD1^bcd^* | 16 | Ast | 6.28E-08 | NA | *CCSER1^abd^* | 4 | In | 6.26E-08 | 1.06E-02 |
| *GFAP^bcd^* | 17 | Ast | 7.24E-08 | 6.97E-03 | *GPNMB^abcd^* | 7 | In | 2.33E-07 | NA |
| *C1QL1^bcd^* | 17 | Ast | 6.32E-17 | 1.29E-02 | *NME3^bcd^* | 16 | In | 1.63E-07 | 5.23E-05 |
| *LRRC37A^bcd^* | 17 | Ast | 6.23E-12 | 3.64E-02 | *DCAKD^bcd^* | 17 | In | 2.84E-19 | 1.50E-24 |
| *LRRC37A2^bcd^* | 17 | Ast | 1.30E-10 | 1.72E-02 | *PLEKHM1^abcd^* | 17 | In | 1.49E-09 | 2.23E-16 |
| *EIF2S2^d^* | 20 | Ast | 1.00E-06 | NA | *CD38^abcd^* | 4 | Oli | 4.73E-13 | NA |
| *KHK^cd^* | 2 | Mic | 2.26E-06 | NA | *GPRIN3^abd^* | 4 | Oli | 9.75E-07 | NA |
| *TMEM163^ad^* | 2 | Mic | 2.16E-11 | NA | *C16orf91^bcd^* | 16 | Oli | 6.45E-07 | NA |
| *BST1^abcd^* | 4 | Mic | 1.06E-06 | NA | *WNT3^abcd^* | 17 | Oli | 5.32E-08 | 4.25E-02 |
| *FAM13A^bd^* | 4 | Mic | 5.01E-09 | 3.59E-36 | *GPR171^d^* | 3 | OPCs | 2.60E-07 | NA |
| *ZSWIM6^bd^* | 5 | Mic | 1.30E-08 | NA | *NELFA^bd^* | 4 | OPCs | 2.36E-06 | NA |
| *GPNMB^abcd^* | 7 | Mic | 2.41E-08 | NA | *FAM13A^bd^* | 4 | OPCs | 1.17E-09 | 3.45E-06 |
| *SPSB3^bcd^* | 16 | Mic | 1.55E-07 | 1.82E-02 | *CCSER1^abd^* | 4 | OPCs | 6.55E-07 | 1.67E-08 |
| *RPRML^bcd^* | 17 | Mic | 2.21E-16 | 5.03E-10 | *FAHD1^bcd^* | 16 | OPCs | 1.26E-07 | NA |
| *GPR171^d^* | 3 | Ex | 1.88E-06 | NA | *TOX3^d^* | 16 | OPCs | 1.36E-06 | NA |
| *FGFRL1^bd^* | 4 | Ex | 3.01E-15 | NA | *NCOR1^bcd^* | 17 | OPCs | 2.36E-06 | NA |
| *CD38^abcd^* | 4 | Ex | 4.41E-10 | NA | *RPRML^bcd^* | 17 | OPCs | 3.22E-20 | 6.35E-03 |
| *GPNMB^abcd^* | 7 | Ex | 7.60E-08 | NA |  | | | | |

- a. Listed as associated with PD traits in GWAS Catalog.
- b. Supported by a significant bulk TWAS-O risk gene by the expanded-cis-window overlap comparison.
- c. Supported by a significant PWAS-O risk gene by the expanded-cis-window overlap comparison.
- d. Independent gene locus, defined by GIFT p-value $\leq0.05$ or as a significant isolated candidate not included in a multigene GIFT analysis.

**Table 3: Significant bulk PWAS-O risk genes of PD in DLPFC.** The table lists 17 protein coding genes meeting the retained significance definition of genomic-control-adjusted $(\lambda\approx1.184)$ omnibus p-values followed by Benjamini-Hochberg correction at $q \leq0.05$.

| **Gene Name** | **CHR** | **Start Position** | **PWAS-O P_ACAT_** | **GC-adjusted BH FDR** | **GIFT P-value** |
| --- | --- | --- | --- | --- | --- |
| *SNX17^d^* | 2 | 27370496 | 2.05E-06 | 6.73E-03 | NA |
| *EXOSC7^d^* | 3 | 44976249 | 2.78E-07 | 1.58E-03 | NA |
| *PDHB^b^* | 3 | 58427630 | 1.95E-05 | 3.75E-02 | 9.37E-01 |
| *FAM107A^bd^* | 3 | 58564112 | 3.05E-05 | 3.76E-02 | 8.64E-16 |
| *CD38^abd^* | 4 | 15778308 | 1.50E-16 | 5.19E-11 | NA |
| *GPNMB^abd^* | 7 | 23246697 | 2.43E-09 | 4.96E-05 | NA |
| *CTSB^abd^* | 8 | 11842525 | 2.41E-05 | 3.76E-02 | NA |
| *SEC23IP^ad^* | 10 | 119892573 | 4.03E-05 | 4.49E-02 | NA |
| *VPS26B^d^* | 11 | 134224605 | 1.45E-07 | 1.07E-03 | NA |
| *RPS2^b^* | 16 | 1962061 | 1.68E-05 | 3.63E-02 | 1.73E-01 |
| *RAB26^b^* | 16 | 2148144 | 2.62E-05 | 3.76E-02 | 7.69E-01 |
| *MLST8^b^* | 16 | 2205177 | 3.02E-05 | 3.76E-02 | 8.01E-01 |
| *TTC19^bd^* | 17 | 15999380 | 2.71E-05 | 3.76E-02 | NA |
| *GFAP^b^* | 17 | 44905626 | 7.84E-07 | 3.35E-03 | 1.99E-01 |
| *NMT1^bd^* | 17 | 45061299 | 4.93E-20 | 1.75E-13 | 2.83E-10 |
| *HEXIM1^b^* | 17 | 45147317 | 5.62E-09 | 8.10E-05 | 2.77E-01 |
| *GOSR2^bd^* | 17 | 46923120 | 1.20E-16 | 5.19E-11 | 3.34E-08 |

- a. Listed as associated with PD traits in GWAS Catalog.
- b. Supported by a significant bulk TWAS-O risk gene by expanded-cis-window comparison.
- d. Independent gene locus, defined by GIFT p-value $\leq0.05$ or as a significant isolated candidate not included in a multigene GIFT analysis.

**Table 4: Summary of cell-type-aware TWAS-O risk genes of PD.** Cross-omics columns report expanded-cis-window support among the independent signals. The support columns report expanded-cis-window support among independent cell-type-aware signals. Support was defined as overlap on the same chromosome after expanding both gene intervals by $\pm$1 Mb.

| **Cell Type** | **Significant TWAS-O genes** | **Genes reported in GWAS Catalog** | **Independent gene loci** | **Independent gene loci supported by bulk TWAS-O locus in DLPFC** | **Independent gene loci supported by bulk PWAS-O locus in DLPFC** | | **Independent gene loci supported by bulk TWAS-O locus in substantia nigra** |
| --- | --- | --- | --- | --- | --- | --- | --- |
| Astrocytes | 21 | 5 | 10 | 7/10 (70.0%) | 6/10 (60.0%) | 7/10 (70.0%) | |
| Microglia | 24 | 8 | 8 | 6/8 (75.0%) | 5/8 (62.5%) | 5/8 (62.5%) | |
| Excitatory neurons | 25 | 6 | 5 | 4/5 (80.0%) | 3/5 (60.0%) | 3/5 (60.0%) | |
| Inhibitory neurons | 19 | 9 | 8 | 6/8 (75.0%) | 6/8 (75.0%) | 5/8 (62.5%) | |
| Oligodendrocytes | 21 | 9 | 4 | 4/4 (100.0%) | 3/4 (75.0%) | 3/4 (75.0%) | |
| Oligodendrocyte precursor cells (OPCs) | 23 | 6 | 8 | 6/8 (75.0%) | 3/8 (37.5%) | 6/8 (75.0%) | |
