## Supplementary Figures for "Integrating dorsolateral prefrontal cortex multi-omics and GWAS summary data enhances gene-mapping of Parkinson’s disease"

**Supplementary Information**

**Supplementary Data 1-12 are provided in the spreadsheet.**

**Supplementary Data 1. Significant bulk DLPFC TWAS-O results for Parkinson’s disease.** Omnibus and component-method P values, GIFT results, annotations, and independence status are reported for 39 significant genes, including 19 independent signals.

**Supplementary Data 2. Significant cell-type-aware TWAS-O results in astrocytes.** Omnibus and component-method P values, GIFT results, annotations, and independence status are reported for 21 significant associations, including 10 independent signals.

**Supplementary Data 3. Significant cell-type-aware TWAS-O results in microglia.** Omnibus and component-method P values, GIFT results, annotations, and independence status are reported for 24 significant associations, including eight independent signals.

**Supplementary Data 4. Significant cell-type-aware TWAS-O results in excitatory neurons.** Omnibus and component-method P values, GIFT results, annotations, and independence status are reported for 25 significant associations, including five independent signals.

**Supplementary Data 5. Significant cell-type-aware TWAS-O results in inhibitory neurons.** Omnibus and component-method P values, GIFT results, annotations, and independence status are reported for 19 significant associations, including eight independent signals.

**Supplementary Data 6. Significant cell-type-aware TWAS-O results in oligodendrocytes.** Omnibus and component-method P values, GIFT results, annotations, and independence status are reported for 21 significant associations, including four independent signals.

**Supplementary Data 7. Significant cell-type-aware TWAS-O results in oligodendrocyte precursor cells (OPCs).** Omnibus and component-method P values, GIFT results, annotations, and independence status are reported for 23 significant associations, including eight independent signals.

**Supplementary Data 8. Significant bulk DLPFC PWAS-O results for Parkinson’s disease.** Raw and genomic-control-adjusted omnibus P values, FDR q values, component-method P values, GIFT results, annotations, and independence status are reported for 17 significant proteins, including 11 independent signals.

**Supplementary Data 9. Significant bulk TWAS-O results in substantia nigra.** Omnibus and component-method P values, GIFT results, and independence status are reported for 74 significant genes, including 27 independent signals.

**Supplementary Data 10. Internal support from the xWAS-O risk genes of global parkinsonian score and PD-related neuropathologic phenotype in ROS/MAP.** Nominal xWAS p-values for global parkinsonian score (n = 1,187) and the PD-related neuropathologic phenotype (n = 1,194), together with matched component-direction comparisons, are reported.

**Supplementary Data 11. Gene-set enrichment results from pathDIP.** Pathway-enrichment statistics and contributing genes are reported separately for bulk TWAS-O, combined cell-type-aware TWAS-O, and bulk PWAS-O findings.

**Supplementary Data 12. Pairwise sample overlap across molecular-reference and ROS/MAP phenotype cohorts.** Sample sizes, numbers of overlapping participants, and pairwise overlap percentages are reported.

**Supplementary Figures**

**Fig. S1. Quantile-quantile plots of bulk DLPFC TWAS results.**

Results are shown for (A) TWAS-O, (B) TIGAR/DPR, (C) PrediXcan/elastic net, and (D) FUSION/BestModel; $\lambda_{GC}$ = 1.149, 1.206, 1.231, and 1.229, respectively. The dashed diagonal line represents the null expectation.

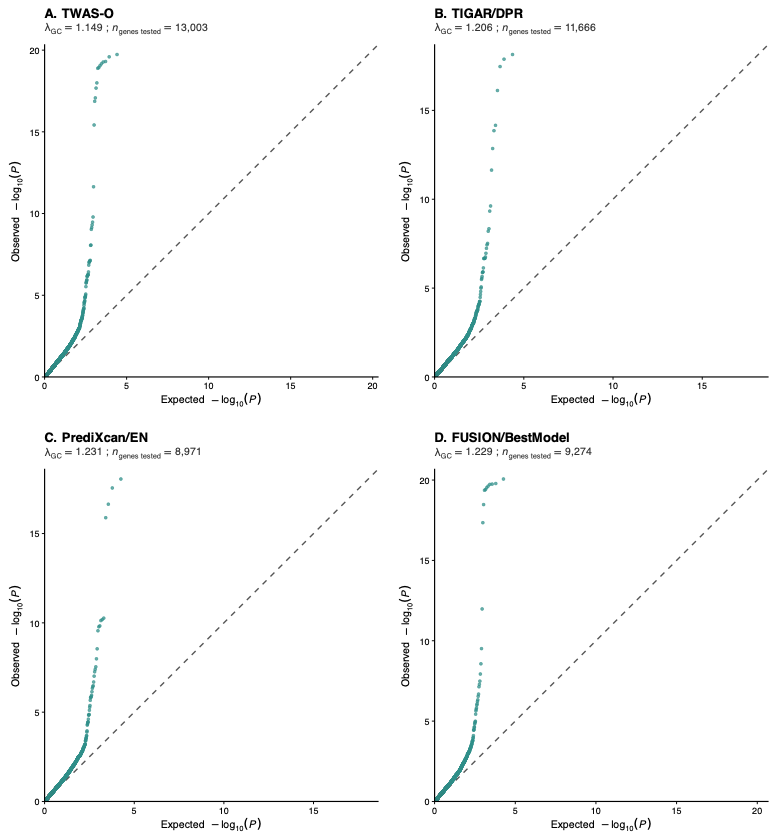

**Fig. S2. Quantile-quantile plots of cell-type-aware DLPFC TWAS results in astrocytes.**

Results are shown for (A) TWAS-O, (B) TIGAR/DPR, (C) PrediXcan/elastic net, and (D) FUSION/BestModel; $\lambda_{GC}$ = 1.158, 1.159, 1.208, and 1.153, respectively. The dashed diagonal line represents the null expectation.

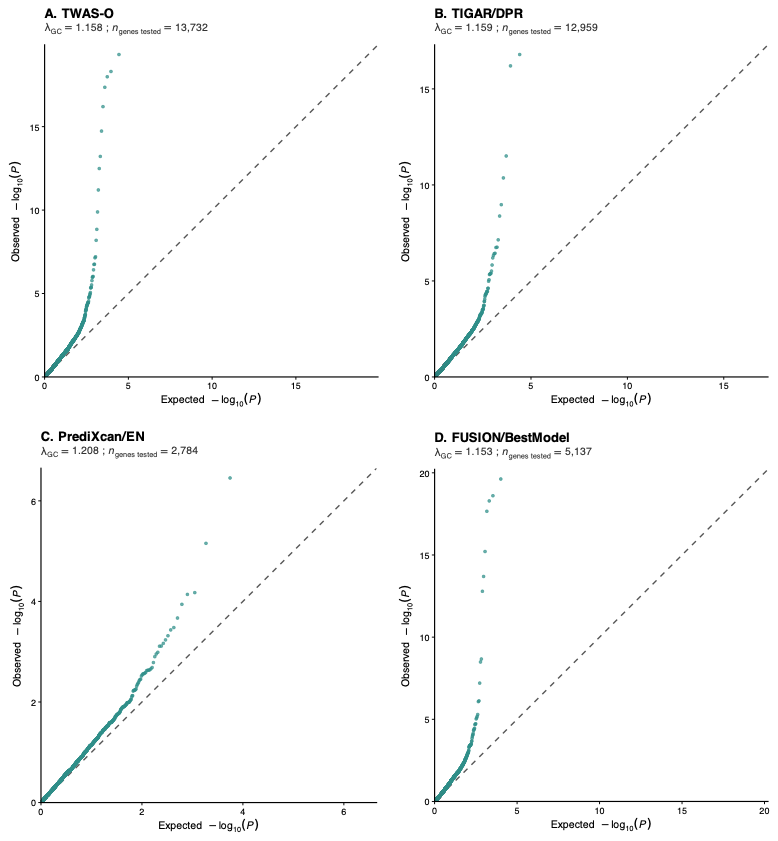

**Fig. S3. Quantile-quantile plots of cell-type-aware DLPFC TWAS results in microglia.**

Results are shown for (A) TWAS-O, (B) TIGAR/DPR, (C) PrediXcan/elastic net, and (D) FUSION/BestModel; $\lambda_{GC}$ = 1.159, 1.195, 1.121, and 1.211, respectively. The dashed diagonal line represents the null expectation.

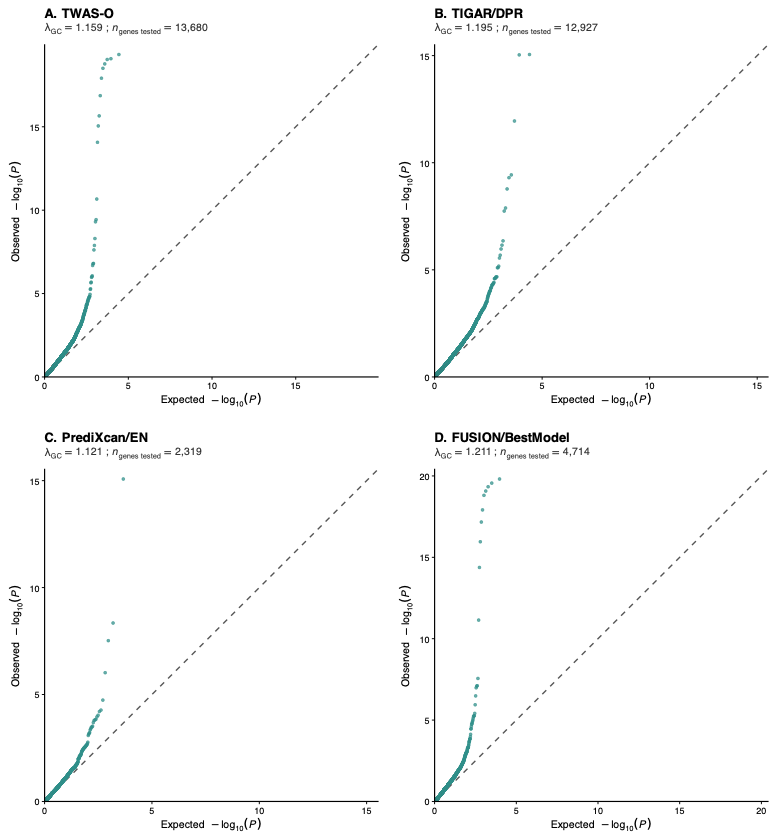

**Fig. S4. Quantile-quantile plots of cell-type-aware TWAS results in excitatory neurons.**

Results are shown for (A) TWAS-O, (B) TIGAR/DPR, (C) PrediXcan/elastic net, and (D) FUSION/BestModel; $\lambda_{GC}$ = 1.155, 1.164, 1.203, and 1.189, respectively. The dashed diagonal line represents the null expectation.

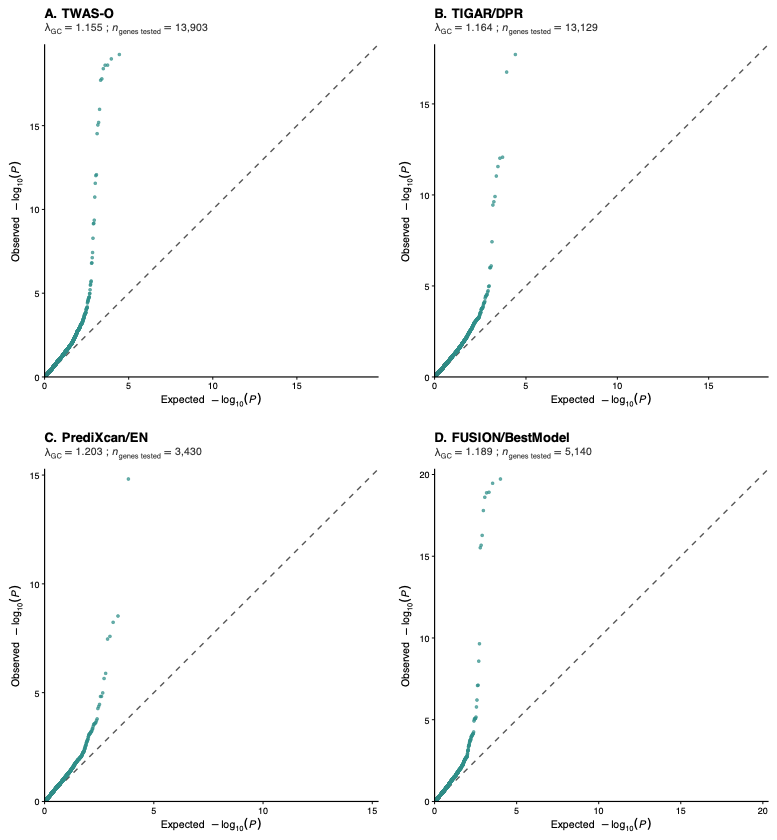

**Fig. S5. Quantile-quantile plots of cell-type-aware TWAS results in inhibitory neurons.**

Results are shown for (A) TWAS-O, (B) TIGAR/DPR, (C) PrediXcan/elastic net, and (D) FUSION/BestModel; $\lambda_{GC}$ = 1.113, 1.122, 1.181, and 1.205, respectively. The dashed diagonal line represents the null expectation.

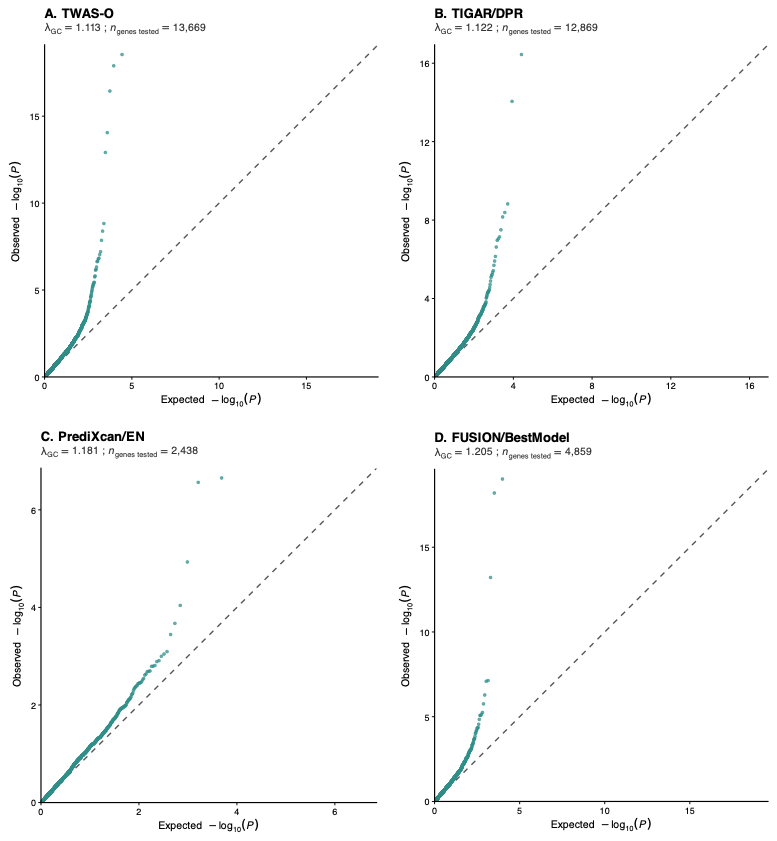

**Fig. S6. Quantile-quantile plots of cell-type-aware TWAS results in oligodendrocytes.**

Results are shown for (A) TWAS-O, (B) TIGAR/DPR, (C) PrediXcan/elastic net, and (D) FUSION/BestModel; $\lambda_{GC}$ = 1.190, 1.204, 1.220, and 1.229, respectively. The dashed diagonal line represents the null expectation.

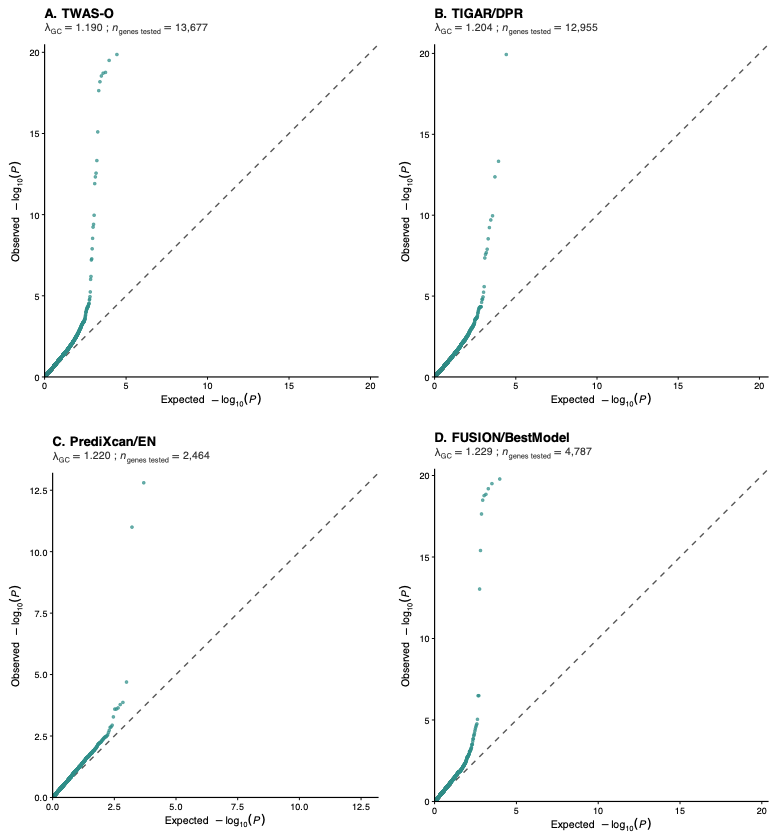

**Fig. S7. Quantile-quantile plots of cell-type-aware TWAS results in oligodendrocyte precursor cells (OPCs).**

Results are shown for (A) TWAS-O, (B) TIGAR/DPR, (C) PrediXcan/elastic net, and (D) FUSION/BestModel; $\lambda_{GC}$ = 1.157, 1.181, 1.235, and 1.254, respectively. The dashed diagonal line represents the null expectation.

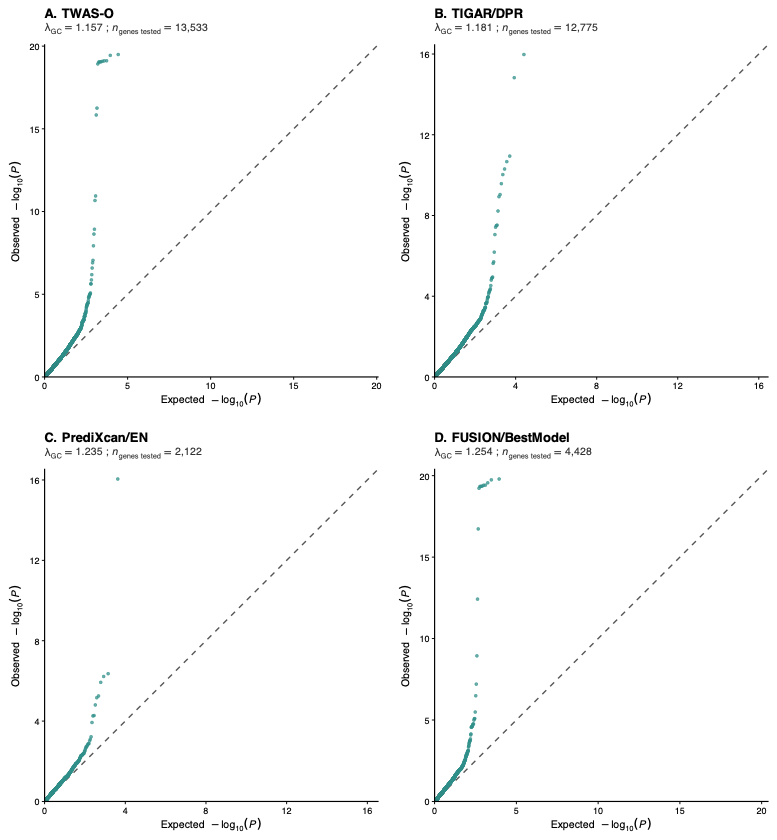

**Fig. S8. Quantile–quantile plots of bulk DLPFC proteome-wide association results.**

Unadjusted results are shown for (A) PWAS-O, (B) TIGAR/DPR, (C) PrediXcan/elastic net, and (D) FUSION/BestModel; $\lambda_{GC}$ = 1.184, 1.195, 1.330, and 1.260, respectively. Genomic control reduced the omnibus PWAS-O $\lambda_{GC}$ to 1.000 before FDR correction.

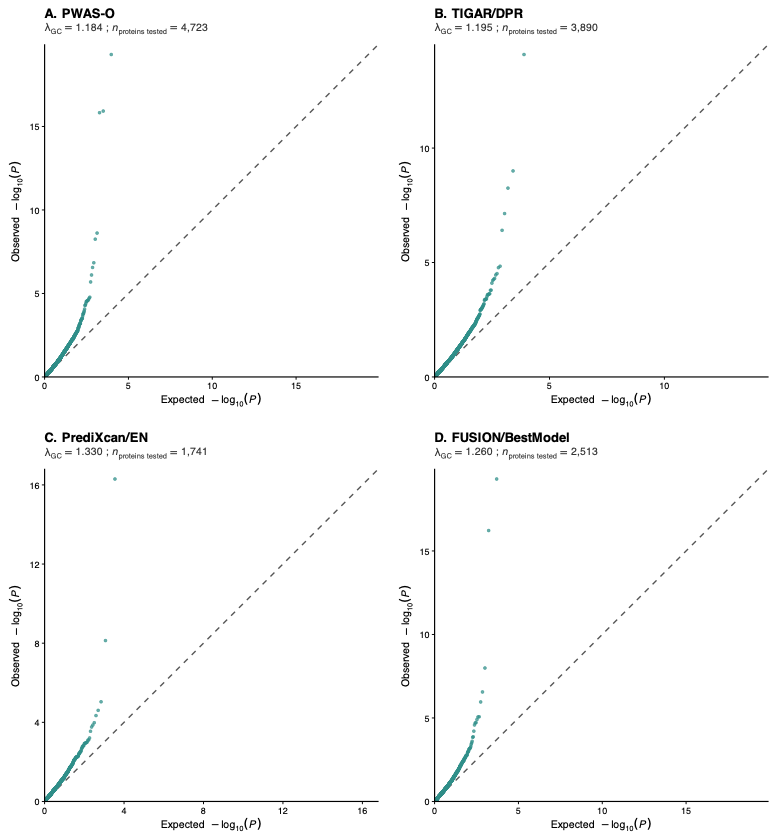

**Fig. S9. GTEx v10 substantia nigra TWAS-O results for Parkinson's disease.**

The Manhattan plot shows -log_10_(TWAS-O p-value) across chromosomes 1-22. The dashed horizontal line indicates the transcriptome-wide significant threshold of $p_{ACAT}=2.5\times{10}^{-6}$. Independent signals are shown in red and labeled; the remaining significant associations are shown in orange. Independent signals were defined by GIFT $p\leq0.05$ or as significant isolated candidates not included in a multigene GIFT analysis.

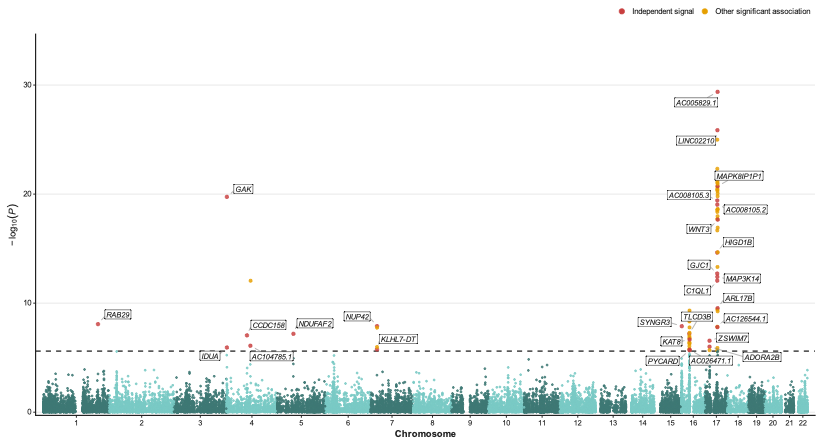

**Fig. S10. GIFT locus plots for multigene GTEx v10 substantia nigra TWAS-O candidate regions.**

Gray circles, blue squares, and red diamonds represent GWAS, TWAS-O, and GIFT results, respectively; the dashed line marks GIFT $P=0.05$, and prioritized independent signals are labeled.

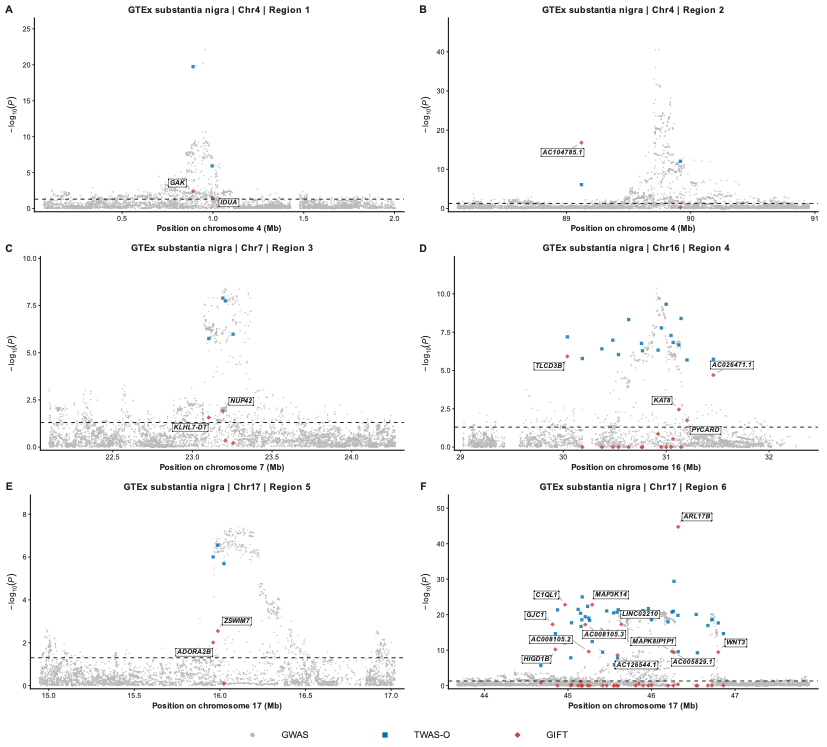

**Fig. S11. GIFT locus plots for multigene bulk DLPFC TWAS-O candidate regions.**

Gray circles, blue squares, and red diamonds represent GWAS, TWAS-O, and GIFT results, respectively; the dashed line marks GIFT $P=0.05$, and prioritized independent signals are labeled.

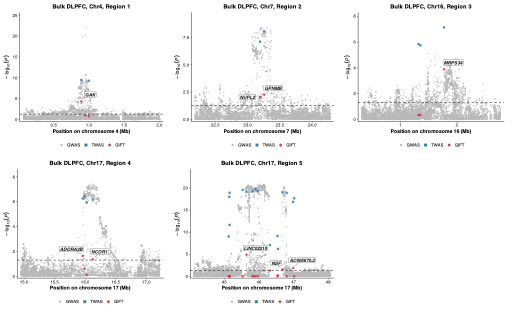

**Fig. S12. GIFT locus plots for multigene cell-type-aware TWAS-O candidate regions in oligodendrocytes and oligodendrocyte precursor cells.**

Gray circles, blue squares, and red diamonds represent GWAS, TWAS-O, and GIFT results, respectively; the dashed line marks GIFT $P=0.05$, and prioritized independent signals are labeled.

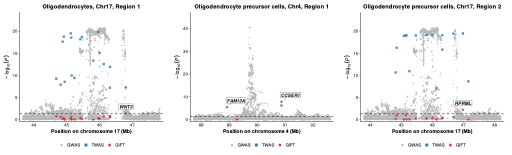

**Fig. S13. GIFT locus plots for multigene bulk DLPFC PWAS-O candidate regions.**

Gray circles, blue squares, and red diamonds represent GWAS, PWAS-O, and GIFT results, respectively; the dashed line marks GIFT $P=0.05$, and prioritized independent signals are labeled.

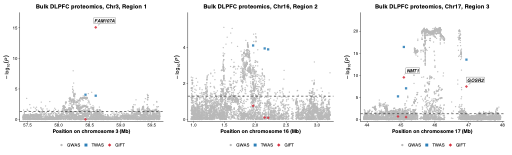

**Fig. S14. Exact-gene overlap among significant DLPFC bulk TWAS-O,**

**cell-type-aware TWAS-O, DLPFC bulk PWAS-O, and GTEx v10 substantia nigra**

**TWAS-O findings.**

Counts represent unique genes, and intersections are based on exact gene-symbol matches across the four analyses.

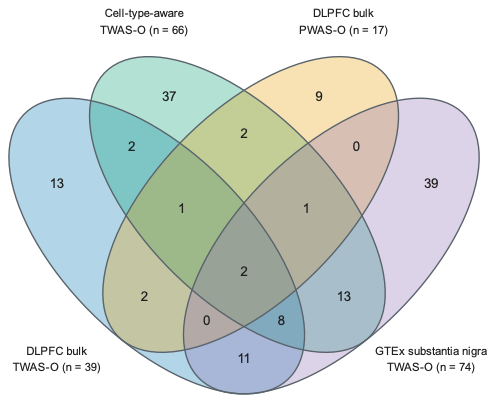

**Fig. S15. Pairwise Jaccard similarity of significant DLPFC TWAS-O and PWAS-O gene sets.**

Pairwise Jaccard indices were calculated among the significant gene sets from DLPFC bulk TWAS-O, DLPFC bulk PWAS-O, the six DLPFC cell-type-aware TWAS-O panels, and GTEx v10 substantia nigra TWAS-O. For gene sets A and B, the Jaccard index was defined as $\left| A\cap B \right|/|A\cup B|$, with only exact gene-symbol matches counted as shared. TWAS-O gene sets were defined using $p_{ACAT}\leq2.5\times{10}^{-6}$, and the PWAS-O gene set was defined using the genomic-control-corrected Benjamini–Hochberg FDR threshold of $q\leq0.05$. All significant genes were included irrespective of GIFT prioritization; diagonal values equal 1.

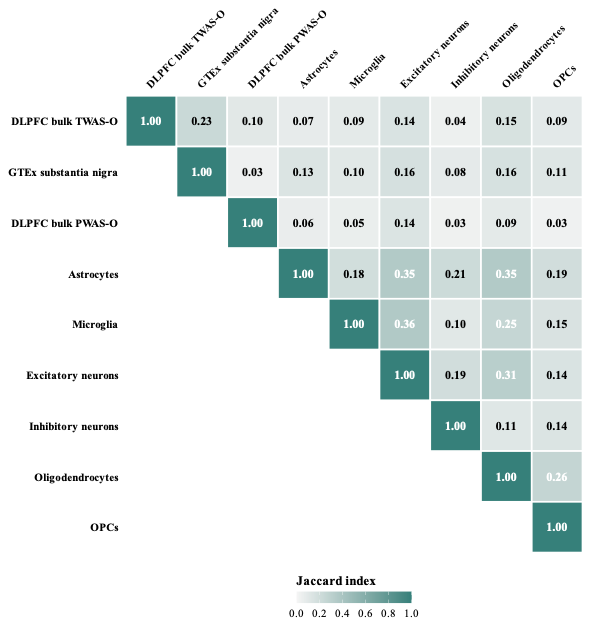

**Fig. S16. Summary of significant and independent genes identified across four molecular-reference analyses.**

(A) Numbers of unique significant and independent genes identified by DLPFC bulk TWAS-O, GTEx v10 substantia nigra TWAS-O, DLPFC cell-type-aware TWAS-O, and DLPFC bulk PWAS-O. (B) Pairwise exact-gene overlaps among the four significant-gene sets. Pairwise counts are not mutually exclusive. Two genes, *GPNMB* and *HEXIM1*, were shared across all four analyses.

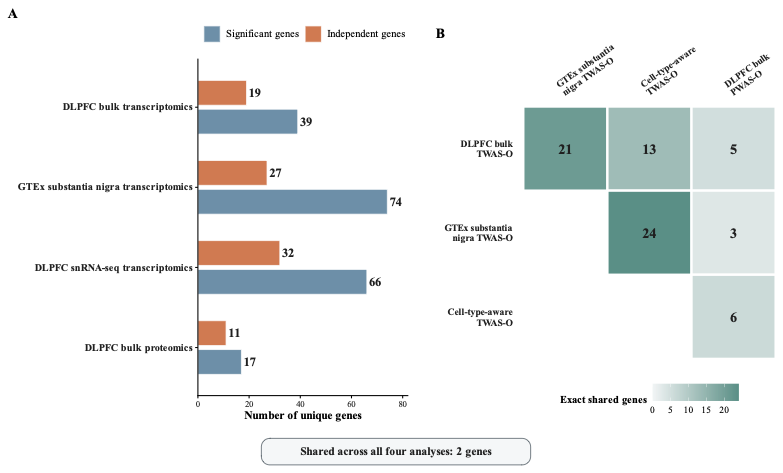
